# Deep learning representations and proteome-wide Mendelian randomization identify causal mediators of myocardial fibrosis

**DOI:** 10.64898/2025.12.13.25342200

**Authors:** Shriya G Reddy, Fang Cao, Roger Xia, Shaun Loong, Ethan Chen, Kirsten Steffner, Jack W O’Sullivan, Francois Haddad, Roger Foo, Victoria N. Parikh, Matthew T. Wheeler, Euan A. Ashley, Bruna Gomes

## Abstract

Cardiac fibrosis is a central pathological process in heart failure, yet the molecular mechanisms governing its spatial organization remain poorly defined. We developed an artificial intelligence (AI)-based phenotyping approach to decode the spatial organization of cardiac fibrosis from routine cardiac magnetic resonance imaging (MRI). We applied convolutional variational autoencoders (VAEs) and distributional metrics to native T1 maps from 50,239 UK Biobank participants. VAE-derived features predicted mortality with greater precision than standard T1 measures (C-index 0.614 vs. 0.547; likelihood ratio p = 2.9×10^-3^), identifying spatial fibrosis patterns as independent prognostic indicators.

Through genome-wide association studies, we identified genetic loci underlying T1 distribution metrics, implicating oxidative stress pathways (SOD2, GSS) and calcium signaling (CAMK2D, CALU). Pathway enrichment revealed distinct biological processes: T1 distributions reflected metabolic and coagulation activity, while spatial VAE dimensions reflected extracellular matrix organization and complement regulation.

Mendelian randomization identified cathepsin S (CTSS) and extracellular matrix protein 1 (ECM1) as causal mediators with near-certain colocalization evidence (PP.H4>0.88), validated in an independent Icelandic cohort (n=35,559). FKBPL demonstrated causal effects on both T1 distributional and spatial features. Published preclinical studies show CTSS inhibition reduces collagen deposition and ventricular stiffening, ECM1 stabilizes extracellular matrix and prevents fibrosis, and FKBPL peptides attenuate fibroblast activation. These findings highlight tractable pathways for therapeutic modulation of myocardial fibrosis.

## Introduction

Heart failure remains a major global health challenge, affecting over 64 million people worldwide and accounting for nearly 14% of deaths in the United States^1,2^. Cardiac fibrosis, marked by extracellular matrix accumulation and myocardial stiffening, emerges across diverse heart diseases and independently predicts up to a five-fold increase in sudden cardiac death or life-threatening arrhythmias^3–5^ **(Figure 1)**. Despite its prognostic importance, no current therapies effectively reverse established fibrosis, and targeted antifibrotic agents tested to date have not meaningfully improved clinical outcomes^6–10^. Addressing this therapeutic gap requires accurate quantitative biomarkers and large-scale data to link imaging traits to their genetic and molecular determinants.

**Figure 1:**
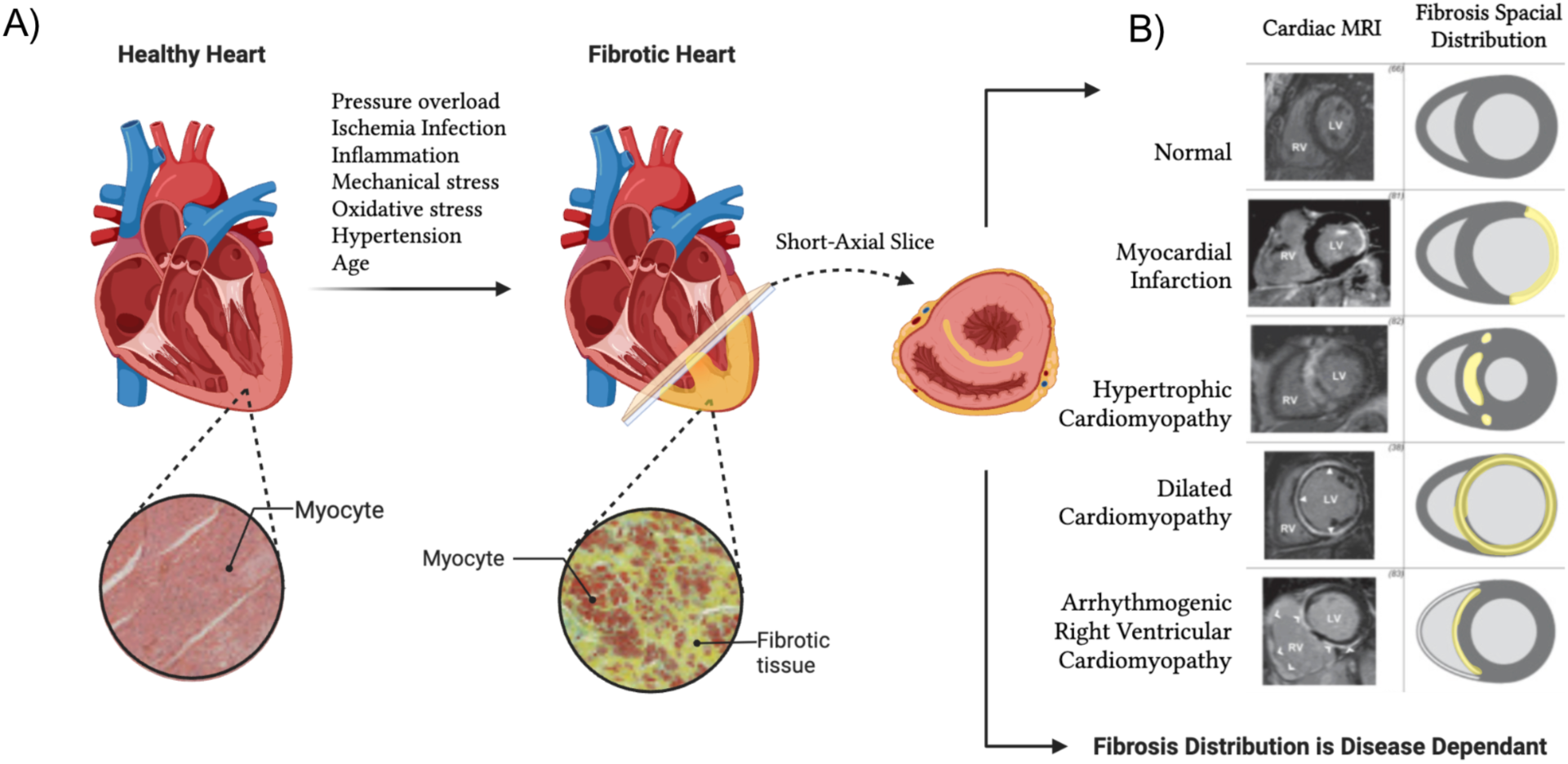
Fibrosis Development in Heart Disease Pathophysiology and Spatial Distribution Patterns. **(A)** Progression from a healthy heart to a fibrotic heart due to various stressors and cellular-level changes in myocyte structure and collagen deposition. **(B)** Characteristic spatial patterns of fibrosis distribution across different cardiac pathologies illustrate how normal, myocardial infarction, hypertrophic cardiomyopathy, dilated cardiomyopathy, and arrhythmogenic right ventricular cardiomyopathy each present distinct fibrotic patterns that can be detected on cardiac MRI.

Cardiac magnetic resonance T1 mapping provides a non-invasive marker of diffuse myocardial fibrosis, with elevated native T1 values reflecting collagen deposition and increased tissue water content^11–18^. The UK Biobank’s population-scale T1 data (>50,000 participants) enables unprecedented power to dissect fibrosis biology^19^. Prior genome-wide association studies using mean septal T1 identified loci linked to oxidative stress, iron homeostasis, and calcium signaling^20^. However, reducing the full T1 map to a single mean value ignores spatial and distributional information that encodes distinct biological processes.

Artificial intelligence (AI) now enables extraction of high-resolution fibrosis phenotypes from native T1 maps, overcoming the limitations of conventional mean-value metrics. Automated deep learning segmentation of the myocardium allows precise, population-scale extraction of percentile-based scalar metrics, to capture the distribution of T1 values across the myocardium^21,22^. As observed in other organs, the geometry of fibrosis reflects underlying molecular pathways and disease stages^23,24^. To capture these features computationally, we employ variational autoencoders, which condense high-dimensional imaging data into a low-dimensional latent space while retaining anatomical structure^25–29^. By automating segmentation and feature extraction at scale, AI phenotyping transforms T1 maps into these low-dimensional datasets, facilitating integration with genetic and proteomic data for discovery.

At the molecular level, observational proteomic studies have associated circulating proteins with myocardial fibrosis, yet such findings cannot establish causality^30,31^. Mendelian randomization provides a framework to infer causal relationships by using genetic variants as instruments that approximate randomized exposure to different protein levels^32^. Recent applications of proteome-wide MR have successfully identified causal protein-disease relationships in heart failure and other cardiovascular conditions, transforming candidate protein biomarkers into potential therapeutic targets^33,34^.

Leveraging the integration of imaging, genomics, and proteomics in the UK Biobank, this study improves the phenotypic definition of myocardial fibrosis and identifies causal molecular determinants. We derive percentile-based and VAE-derived features from native T1 maps, test their predictive value for mortality, perform genome- and proteome-wide association analyses, and apply the first proteome-wide Mendelian randomization to identify causal circulating mediators of diffuse myocardial fibrosis **(Figure 2)**. Together, these analyses reveal modifiable biological pathways that may inform the development of targeted antifibrotic therapies.

**Figure 2:**
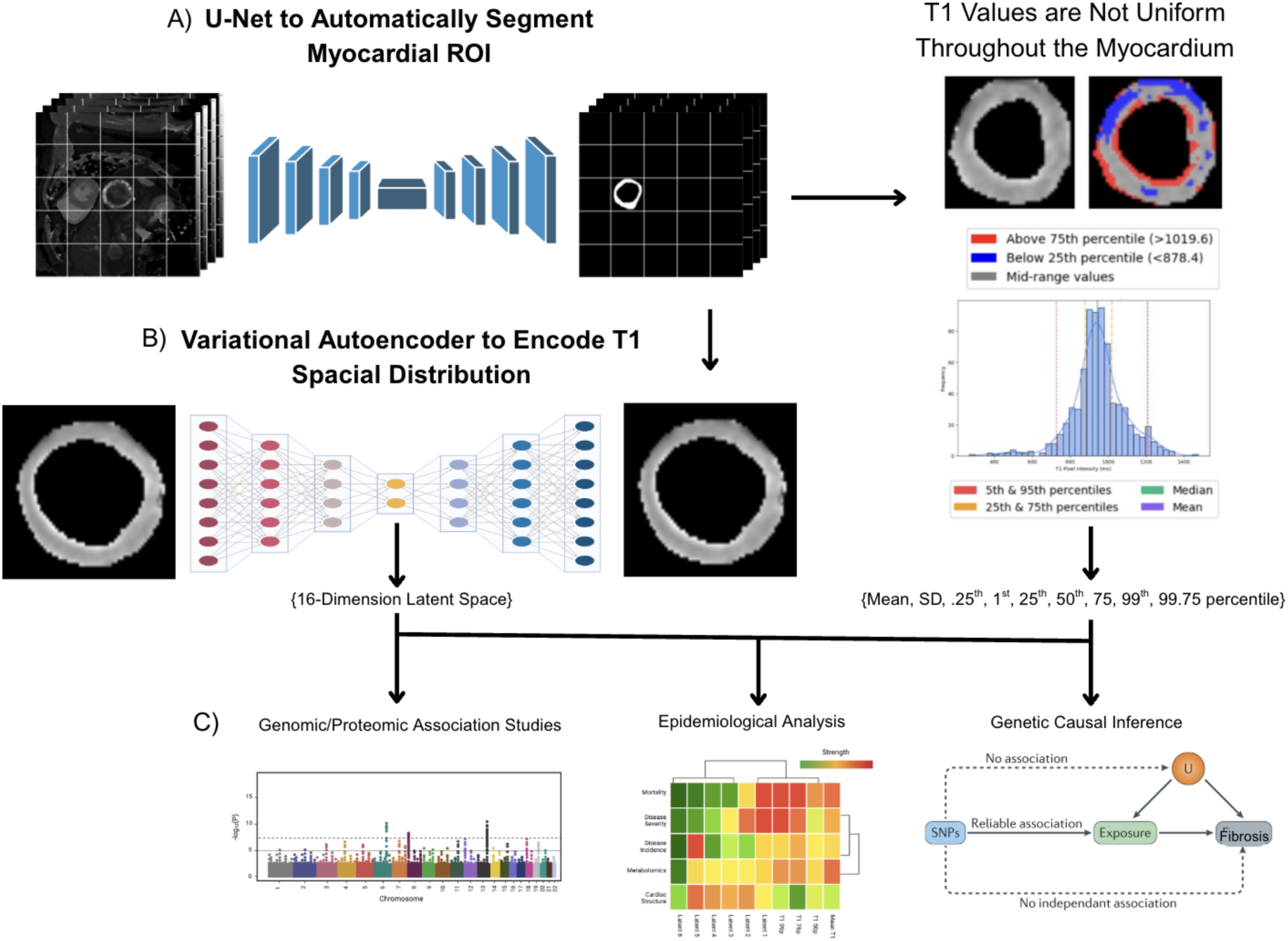
Methodological Framework for Spatial Analysis of Myocardial Fibrosis. Flowchart illustrating the multi-step analytical pipeline developed in this study. **(A)** U-Net segmentation of myocardial regions of interest from native T1-mapping images. **(B)** Extraction of conventional scalar metrics alongside spatial heterogeneity analysis through variational autoencoder encoding of myocardial T1 maps into a 16-dimensional latent space. **(C)** Comprehensive downstream analytical approaches including: 1) genomic/proteomic association studies (GWAS, PWAS, heritability analysis and rare variant burden testing); 2) clinical relevance analysis for phenotype associations, mortality (Kaplan-Meier survival curves, log-rank tests, Cox models) and disease associations (delta rank analysis, disease prevalence across quartiles); and 3) Mendelian randomization to establish causal relationships between protein biomarkers and myocardial fibrosis phenotypes

## Results

### Automated extraction of T1 distribution metrics characterizes myocardial T1 variability at population scale

We developed an automated pipeline to segment a short-axis slice of the left ventricular myocardium **(Figure 3A)** and quantify T1 distributions in 50,239 UK Biobank participants (mean age 64.0±7.7 years, 48.6% male) who underwent native T1 mapping using Shortened Modified Look-Locker Inversion recovery (ShMOLLI)^12,35^. U-Net models achieved robust segmentation performance (Sørensen–Dice = 0.84, 95% CI: 0.83–0.85), with automated measurements closely matching manual annotations (r^2^ = 0.914; **Supplementary Figure 3)**. After quality control, segmentation succeeded in 42,083 participants (83.7% overall).

**Figure 3:**
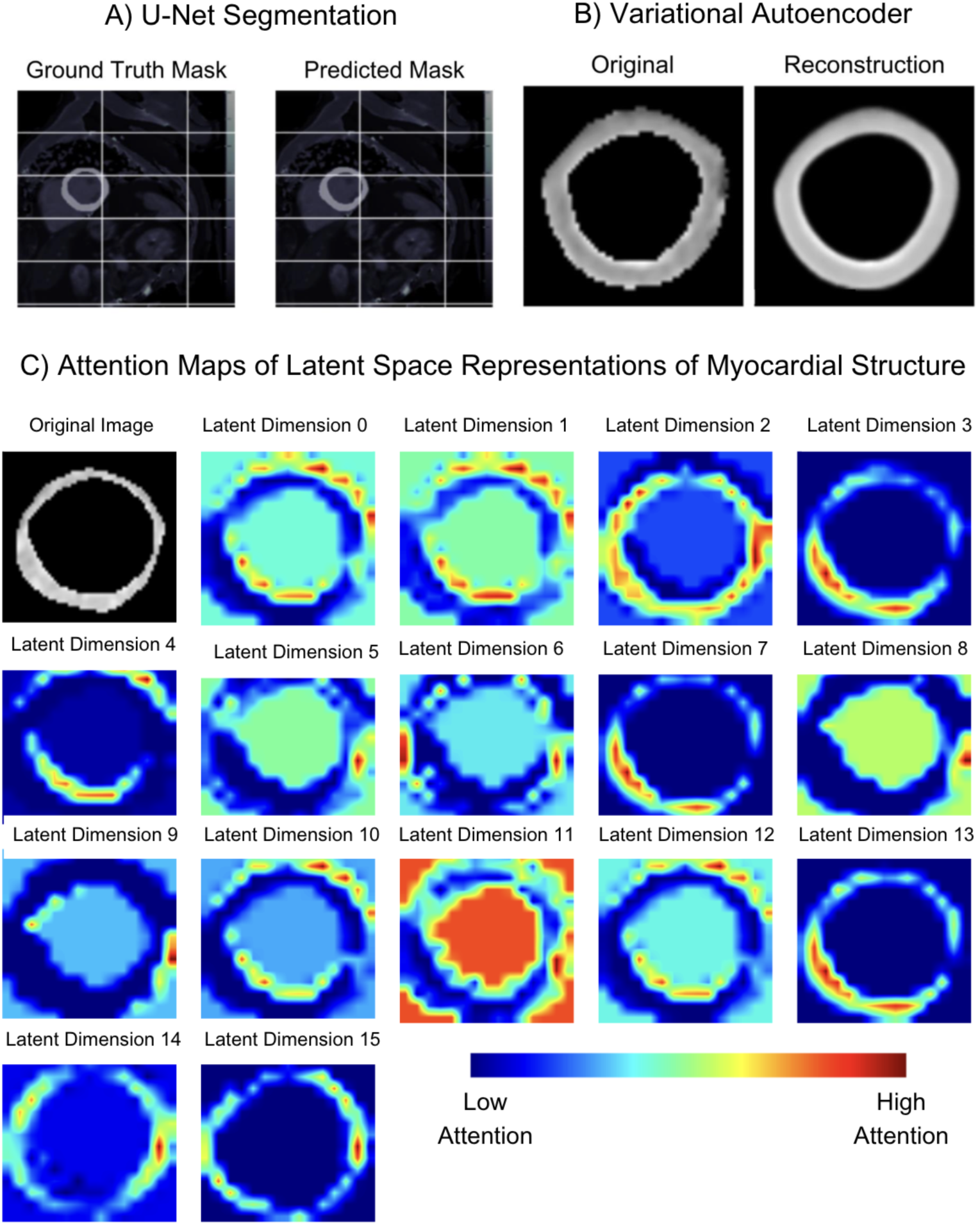
Deep Learning Model Performance and Latent Space Interpretability for Myocardial Analysis. **A)** U-Net segmentation results comparing ground truth masks with model predictions, demonstrating the network’s ability to delineate myocardial boundaries accurately. (**B)** Variational autoencoder reconstruction showing the original myocardial image and its decoded output, illustrating the model’s capacity to learn compressed latent representations that preserve structural information. **(C)** Visualization of gradient-based attention maps for all 16 latent dimensions, showing which regions of the myocardium most strongly influence each dimension. The original image (grayscale) is shown at the top left.

From each T1 map, we extracted “distribution metrics” capturing both central tendency and extremes: percentiles (0.25th, 1st, 25th, 50th, 75th, 99th, 99.75th), mean, and standard deviation **(Table S1)**. All metrics were adjusted for age and sex to account for known physiological variation^36,37^. While these metrics characterize T1 distributions, they do not preserve spatial organization within the myocardium, motivating our second analytical approach.

### Variational autoencoder quantifies myocardial T1 spatial organization through latent dimensions

To capture spatial heterogeneity, we implemented a convolutional variational autoencoder (CVAE) that learns unsupervised representations of regional T1 patterns **(Figure 3B)**. Each latent dimension summarizes a specific spatial pattern as a single numerical value. The CVAE, trained with 16 latent dimensions (optimized through systematic evaluation detailed in **Methods and Supplementary Figure 2**), achieved high reconstruction fidelity on 1,000 validation images: mean squared error (MSE) of 0.0005, peak signal-to-noise ratio (PSNR) of 32.15 dB, structural similarity index (SSIM) of 0.92, and Dice score of 0.92 for myocardial boundary preservation. These metrics validated the model’s ability to capture essential morphological features while maintaining low-dimensional representation.

To interpret what spatial information each latent dimension encoded, we applied gradient-based attention mapping^38^, which quantifies the influence of each image region on a given latent dimension by computing the gradient of that dimension with respect to input pixels **(Table S2, Figure 3C)**. Attention maps were independently reviewed by a level III cardiovascular MRI expert (B.G.) to assign anatomical interpretations. This revealed functional specialization across dimensions: dimension 11 showed strong attention to image background; dimensions 3, 7, and 13 predominantly encoded the interventricular septum; dimensions 5, 6, 8, and 9 focused on specific lateral wall regions; dimensions 14 and 15 captured broader myocardial patterns excluding the septum; and dimensions 0, 1, 10, and 12 represented the superior myocardial boundary. This spatial organization emerged without explicit supervision, suggesting the model learned biologically meaningful features.

### Latent dimensions capture additional prognostic value beyond standard native T1 metrics

To evaluate the clinical relevance of T1 distribution metrics and VAE latent dimensions, we examined their associations with prevalent cardiovascular disease and mortality. We compared participants with prevalent disease **(see Methods for ICD-10 mapping)** to healthy controls using delta rank analysis **(Figure 4D, Supplementary Figure 4)**. Hypertension (n = 5,597) demonstrated widespread associations across T1 distribution metrics (mean T1: delta rank = 0.049, p = 5.7×10^-9^; 75th percentile: delta rank = 0.046, p = 6.7×10^-9^). Ischemic heart disease (n = 1,807) was significantly associated with T1 percentiles (delta rank = 0.059-0.075), with the 25th percentile demonstrating the strongest association (delta rank = 0.075, p = 1.0×10^-9^). Within this group, myocardial infarction history (n = 561) specifically associated with reduced T1 standard deviation (delta rank = -0.106, p = 1.6×10^-5^).

**Figure 4.**
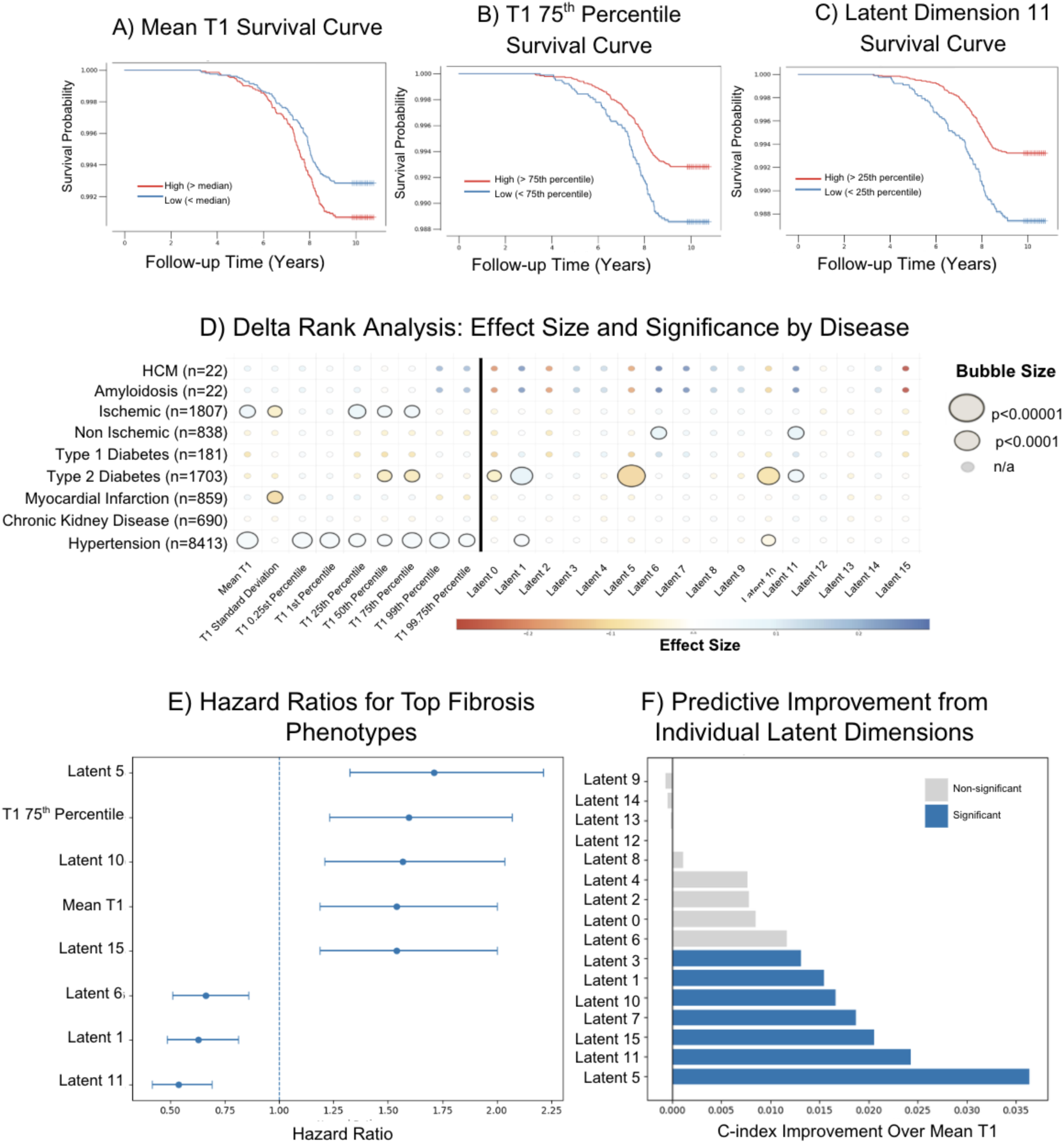
Prognostic and clinical associations of T1 distribution metrics and VAE latent dimensions. Kaplan-Meier survival curves stratified by median for **(A)** mean T1, **(B)** T1 75th percentile, and **(C)** VAE Latent Dimension 11, demonstrating mortality prediction across phenotypes. **(D)** Delta rank analysis showing associations between T1 metrics and VAE latent dimensions with prevalent cardiovascular and metabolic diseases. Bubble size indicates significance; color indicates effect size (red = positive association, blue = negative association). **(E)** Hazard ratios for all-cause mortality across top-performing fibrosis phenotypes. VAE latent dimensions (Latent 5, 10, 11) and T1 75th percentile demonstrate stronger mortality prediction than conventional mean T1. **(F)** Improvement in mortality prediction (C-index) for individual VAE latent dimensions over mean T1.

In contrast, non-ischemic heart disease (n=838) showed weak associations with T1 distribution metrics but significant associations with VAE latent dimensions 6 and 10 (delta rank=0.090, p = 1.5×10^-5^ and 0.092, p = 8.0×10^-6^). Type 2 diabetes (n=1,703) demonstrated associations with both T1 metrics and multiple VAE dimensions. Chi-square analysis across quartiles showed concordant disease associations **(Table S3).** This suggests that spatial patterns, captured by the VAE but not by scalar T1 statistics, may provide value for identifying subtle remodeling in non-ischemic cardiomyopathies.

To evaluate prognostic significance, we performed Kaplan–Meier and Cox proportional-hazards analyses stratified by percentile thresholds for each imaging feature **(Figure 4A-C, Table S4-5)**. Higher T1 percentiles predicted elevated mortality risk (T1 75th percentile HR = 1.60, 95% CI 1.23–2.07; p = 3.8 × 10^-4^), whereas lower percentiles were significantly associated with improved survival (T1 25th percentile HR = 0.66, 95% CI 0.51–0.86; p = 0.0019) **(Table S5)**. Among VAE features, Latent dimension 11 at the 25th percentile yielded the strongest survival separation (log-rank p = 1.1 × 10^-6^; HR = 0.54, 95% CI 0.42–0.69). VAE latent dimensions comprised six of the eight features with the most significant mortality associations **(Figure 4E)**.

Nested Cox models quantified the added predictive value: incorporating all 16 latent dimensions increased the C-index from 0.547 (mean T1 alone) to 0.614 (likelihood ratio p = 2.86×10^-3^), with individual dimensions improving C-index by 0.013-0.036 relative to mean T1 **(Figure 4F, Table S6-7)**. Together, these results suggest that latent dimensions extracted from T1 maps capture complementary biological information and provide measurable gains in disease association and outcome prediction beyond standard T1 metrics

### T1-associated proteins reflect metabolism while spatial patterns capture active tissue remodeling

To determine whether T1 distribution metrics and VAE latent dimensions capture redundant or complementary information, we computed pairwise correlations between all 9 T1 metrics and 16 VAE dimensions after adjusting for age, sex, and BMI **(Table S8)**. Correlations were predominantly weak (|r| < 0.10), with the strongest observed for Latent 2 (r = 0.12–0.16) and Latent 15 (r = 0.09–0.11), dimensions whose attention patterns span the entire myocardium. Region-specific dimensions showed near-zero correlations: Latent 3, 7, and 13, which pay greater attention to the interventricular septum, exhibited |r| < 0.08 with all T1 distribution metrics. This weak correlation structure suggests that spatial patterns captured by the VAE are not simply recapitulations of scalar T1 distributions.

We next examined correlations with cardiac function measures and circulating metabolites. T1 distribution metrics showed weak inverse correlations with LV mass (r = -0.117), wall thickness (r = - 0.124), glucose (r = -0.033), and LDL cholesterol (r = -0.076), consistent with prior UK Biobank observations likely reflecting physiological confounders rather than fibrosis^39,40^. VAE latent dimensions captured stronger associations with LV strain than T1 distribution metrics (dimension 8: r=0.186). All surrogate fibrosis phenotypes demonstrated weak metabolic correlations. (Table S9-12).

To identify circulating protein biomarkers that may more directly reflect the molecular mechanisms underlying these imaging phenotypes, we tested associations with 2,923 plasma proteins (Olink platform) using linear regression adjusted for age and sex. After Bonferroni correction, 80 proteins associated with T1 metrics and 92 with VAE dimensions, with only 36 proteins (45% and 39%, respectively) overlapping between phenotype classes **(Figure 5A-B, Table S13-14)** T1 distribution metrics showed strongest associations with metabolic and endocrine proteins: IGFBP2 (insulin-like growth factor binding protein 2, significant across 7 of 8 T1 metrics, strongest p = 1.0×10^-23^), LEP (leptin, 6 metrics, p = 1.0×10^-20^), PLAT (tissue plasminogen activator, 7 metrics, p = 1.0×10^-16^), and FABP4 (fatty acid binding protein 4, 6 metrics, p = 1.0×10^-15^). VAE latent dimensions showed broader associations with these metabolic proteins (LEP: 10 of 16 dimensions; FABP4: 7 dimensions) but additionally associated with extracellular matrix components including COL4A1 (collagen type IV alpha 1), COL6A3 (collagen type VI alpha 3), and matrix proteoglycans (HSPG2, NCAN, BCAN).

**Figure 5.**
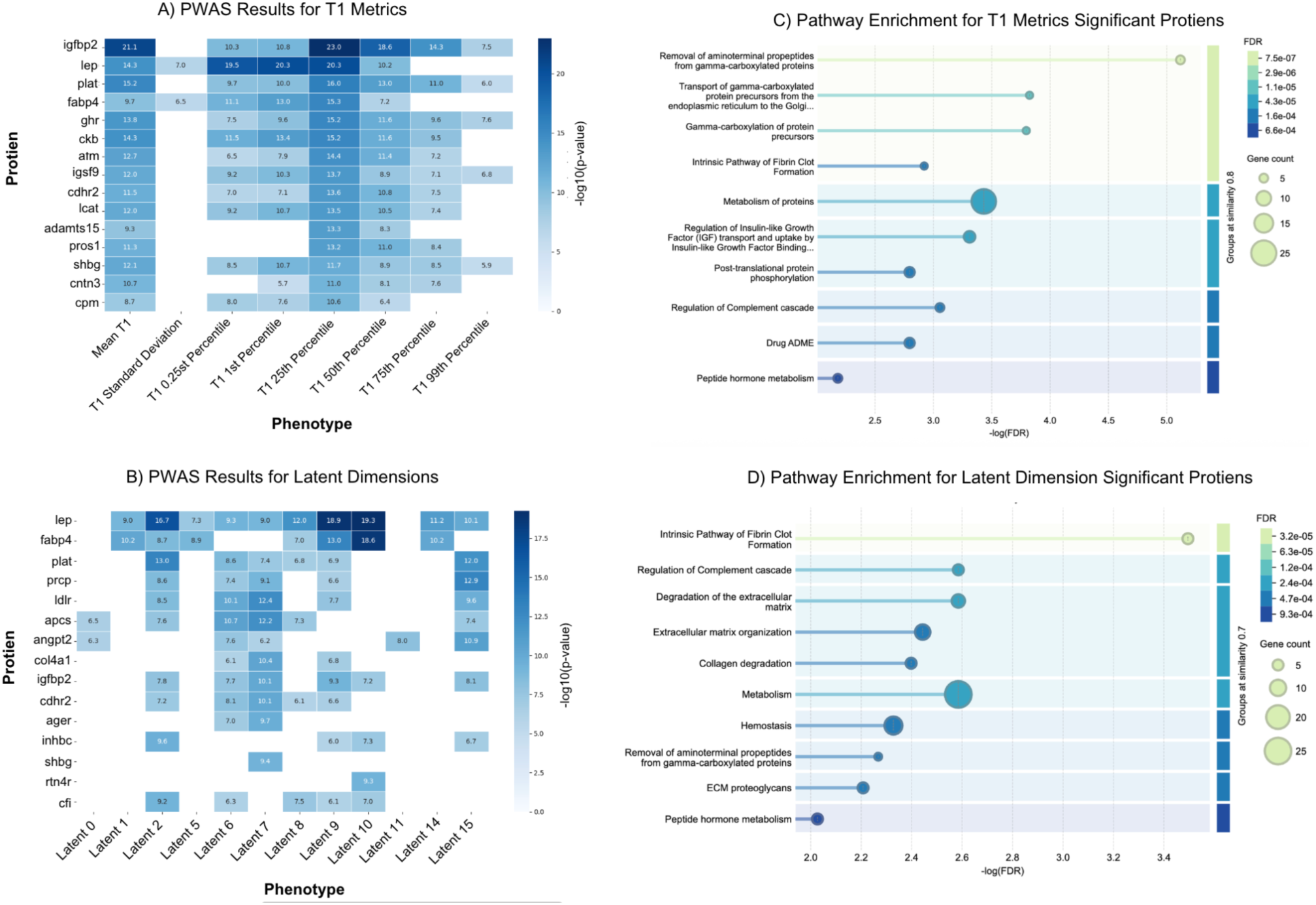
Proteome-wide association study reveals distinct protein signatures for T1 distribution metrics and VAE latent dimensions. **(A)** PWAS results for T1 distribution metrics. Heatmap shows - log10(p-value) for protein associations with mean T1, standard deviation, and percentiles. **(B)** PWAS results for VAE latent dimensions. **(C)** Pathway enrichment for T1 metric-associated proteins and **(D)** for latent dimension-associated proteins. Dominant enrichment for extracellular matrix pathways (ECM degradation, organization, collagen degradation) and complement regulation, contrasting with the metabolic/coagulation signature of T1 metrics.

STRING-based pathway enrichment analysis using the Reactome database revealed fundamentally different biological processes underlying the two phenotype classes **(Figure 5C-D, Table S15-16)**. T1-associated proteins strongly enriched for post-translational protein processing and coagulation: removal of aminoterminal propeptides from gamma-carboxylated proteins (FDR=7.63×10^-6^, 5 genes), gamma-carboxylation of protein precursors (FDR = 1.6×10^-4^, 4 genes), and metabolism of proteins (FDR = 3.7×10^-4^, 24 genes). In contrast, VAE latent dimension-associated proteins enriched for extracellular matrix remodeling: degradation of the extracellular matrix (FDR = 2.6×10^-3^, 7 genes), regulation of complement cascade (FDR = 2.6×10^-3^, 5 genes), and extracellular matrix organization (FDR = 3.6×10^-3^, 9 genes). These results suggest that T1 metrics predominantly reflect systemic metabolic and coagulation factors while VAE dimensions capture extracellular matrix structural organization.

### T1 distribution metrics capture higher genetic variation than latent features

To quantify the heritability of myocardial T1 characteristics attributable to common genetic variants, we estimated SNP-based heritability (h²SNP). LD Score regression (LDSC) was applied to the GWAS summary statistics for all T1 distribution metrics and VAE latent dimensions **(Figure 6E)**.

**Figure 6.**
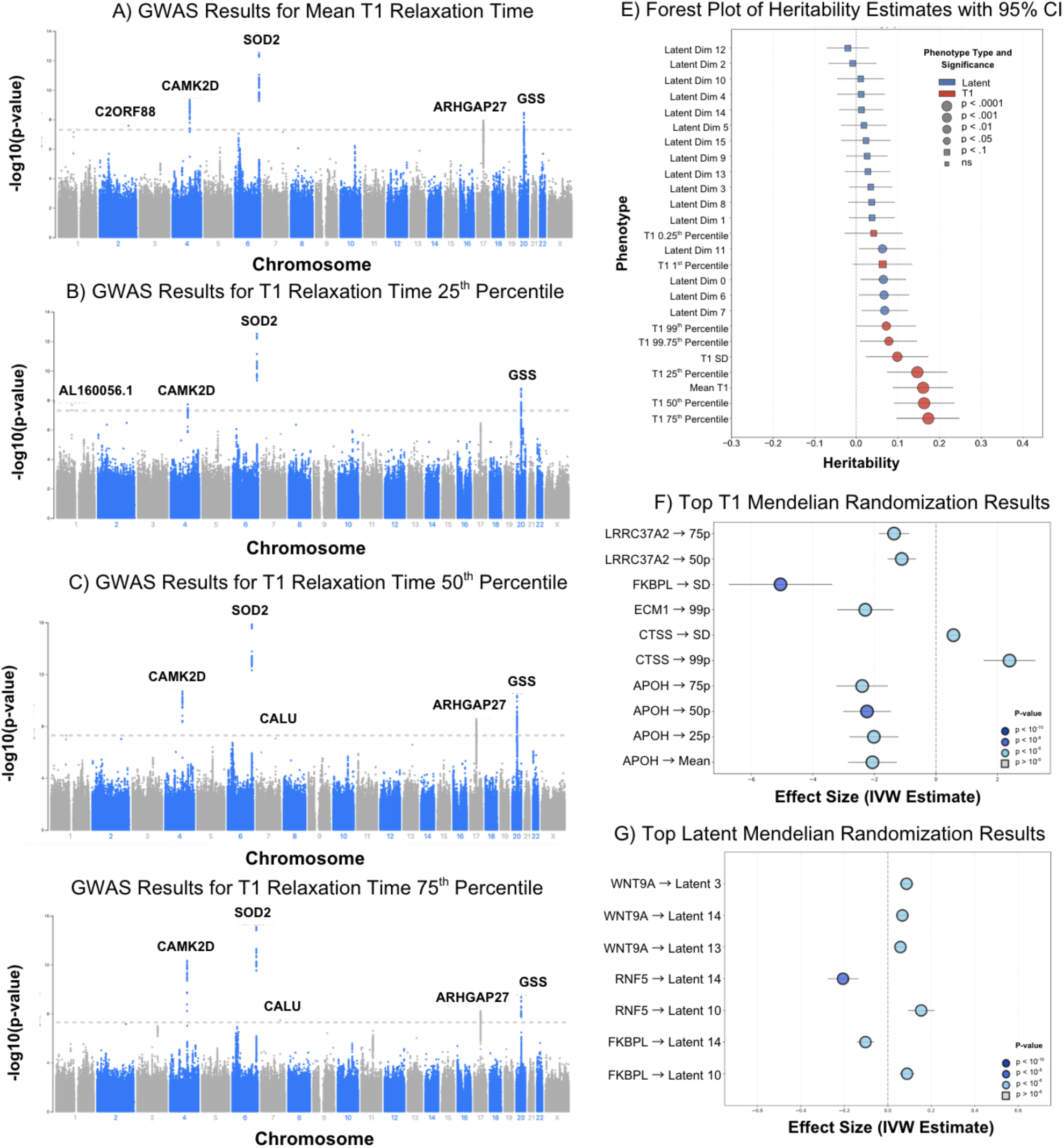
Genome-wide association studies and Mendelian randomization identify genetic determinants and causal proteins. Manhattan plots for GWAS of **(A)** mean T1 and T1 **(B)** 25th, **(C)** 50th, **(D)** 75th percentiles. **(E)** SNP-based heritability estimates (h²) for all T1 metrics and VAE latent dimensions. **(F)** Top Mendelian randomization results for T1 phenotypes and **(G)** VAE latent dimensions.

T1 distribution metrics demonstrated moderate but significant heritability **(Table S17)**, with the 75th percentile showing the highest genetic contribution (h² = 0.173, SE = 0.038, p = 6.3×10^-6^), followed by the 50th percentile (h² = 0.163, SE = 0.037, p = 8.8×10^-6^) and mean T1 (h² = 0.161, SE = 0.037, p = 1.1×10^-5^). This pattern suggests that upper-distribution percentiles capture genetic variability as effectively as or better than conventional mean values used in prior studies^20^. In contrast, VAE latent dimensions showed no significant heritability (**Table S18**), likely reflecting their capture acquired pathological processes such as regional fibrosis, ischemic remodeling, or other forms of spatially-organized cardiac injury that accumulate over the lifespan.

### GWAS identifies oxidative stress and calcium signaling pathways as genetic underpinnings of myocardial T1

Having established that central T1 distribution metrics demonstrate moderate heritability, we performed genome-wide association studies (GWAS) on 35,160 UK Biobank participants with both imaging and genetic data **(Figure 6A-D, Table S19)**.

The strongest signals localized to previously identified SOD2 on chromosome 7 (lead SNP rs2028078, 7:128393950 G/A, intronic, CADD=0.21, p = 7.7×10^-16^ for T1 75th percentile; β = -2.28 ms) and CAMK2D on chromosome 4 (lead SNP 4:114448417 GT/G, intronic, CADD=-0.65, p = 7.1×10^-10^ for mean T1; β = 1.73 ms)^20^. SOD2 encodes mitochondrial superoxide dismutase 2, a key antioxidant enzyme, while CAMK2D encodes calcium/calmodulin-dependent protein kinase critical for excitation-contraction coupling^41,42^. We identified one novel locus at GSS on chromosome 20 (lead SNP 20:33554761 C/T, intronic, CADD=0.53, p = 4.2×10^-10^ for T1 75th percentile; β = -2.06 ms), which encodes glutathione synthetase catalyzing the final step of glutathione biosynthesis, the most abundant antioxidant defense in the cardiovascular system^43,44^.

Three nominally significant loci emerged at CALU on chromosome 17 (lead SNP 17:43475929 G/A, intronic, CADD=-0.02, p = 5.8×10^-9^; β = -1.95 ms), encoding calumenin, a calcium-binding ER chaperone that regulates mitochondrial calcium homeostasis and protects against cardiotoxicity^45^; ARHGAP27 on chromosome 6 (lead SNP 6:160112638 A/G, intronic, CADD=-0.32, p = 5.9×10^-9^), encoding a Rho GTPase-activating protein that regulates cytoskeletal dynamics^46^; and AKAP19 on chromosome 2 (lead SNP 2:190949207, intronic, p = 2.6×10^-8^), encoding a small A-kinase anchoring protein involved in cAMP-PKA signaling^47^. All lead variants were intronic with low CADD scores, indicating likely regulatory rather than protein-damaging effects.

### Mendelian randomization identifies causal effects of circulating proteins on myocardial T1 patterns

To distinguish causal determinants from correlated biomarkers, we performed Mendelian randomization using protein quantitative trait loci (pQTLs) as genetic instruments. After Bonferroni correction, 26 proteins showed causal effects on T1 distribution metrics (53 protein-phenotype pairs) and 27 proteins on VAE latent dimensions (40 protein-phenotype pairs) **(Figure 6F-G, Table S20-21)**.

T1 distribution metrics revealed causal contributions from both immune and structural proteins. Immune proteins including MICB (MHC class I polypeptide-related sequence B, a stress-induced NK cell ligand; β = 0.38, p = 3.0×10^-13^), HLA-E (a non-classical MHC molecule; β = 0.77, p = 1.5×10^-10^), CDSN (corneodesmosin, an epithelial structural protein; β = 0.57, p = 4.8×10^-13^), and C2 (complement component 2; β = 0.98, p = 5.9×10^-11^) all causally increased T1 standard deviation, though colocalization evidence remained weak to modest (PP.H4 = 0.11–0.16). In contrast, extracellular matrix proteins demonstrated both strong causal effects and high colocalization probability **(Table S22-23)**. ECM1 (extracellular matrix protein 1, which stabilizes collagen networks) showed near-certain colocalization (PP.H4=0.958) with causal effects on T1 99th percentile (β = -2.22, p = 8.3×10^-4^), while CTSS (cathepsin S, a lysosomal cysteine protease that degrades ECM components) demonstrated robust effects on both T1 99th percentile (β = 2.47, p = 1.0×10^-4^, PP.H4=0.883) and T1 standard deviation (β = 0.58, p = 1.3×10^-4^, PP.H4=0.728). LRRC37A2 (a leucine-rich repeat protein recently linked to ventricular morphology) exhibited the strongest colocalization evidence (PP.H4>0.999), influencing both T1 75th percentile (β = - 1.31, p = 1.0×10^-4^) and 50th percentile (β = -1.05, p = 1.7×10^-3^). APOH (apolipoprotein H, also known as β2-glycoprotein I) showed protective effects with strong colocalization across multiple percentiles (50th: β = -2.22, p = 1.6×10^-4^, PP.H4=0.819; 75th: β = -2.37, p = 6.3×10^-5^, PP.H4=0.725; mean: β = -2.04, p = 1.9×10^-4^, PP.H4=0.644).

For VAE latent dimensions, Mendelian randomization identified causal effects from immune, metabolic, structural, and epigenetic proteins. Metabolic regulators included DPEP2 (latent 13: β = 0.037, p = 9.7×10^-13^; latent 9: β = -0.023, p = 3.3×10^-10^) and DPY30 (latent 12: β = -0.059, p = 2.6×10^-^¹¹). Structural proteins included DAG1 (latent 13: β = -0.115, p = 5.4×10^-10^) and immune mediators included PTPRC (latent 7: β = 0.032, p = 2.3×10^-9^) and HLA-E (latent 14: β = 0.017, p = 4.2×10^-10^). Among these, three proteins stood out for their effect magnitudes and biological plausibility: FKBPL (FK506-binding protein like, an angiogenesis regulator) showed strong effects on latent dimension 14 (β = -0.10, p = 1.2×10^-7^) and latent dimension 10 (β = 0.089, p = 1.3×10^-7^), RNF5 (an E3 ubiquitin ligase involved in protein degradation) demonstrated the largest effect sizes (latent 14: β = -0.21, p = 4.7×10^-9^; latent 10: β = 0.15, p = 4.8×10^-7^), and WNT9A (a Wnt signaling ligand) exhibited effects across multiple spatial dimensions (latent 3: β = 0.086, p = 4.5×10^-8^; latent 13: β = 0.058, p = 7.7×10^-8^; latent 14: β = 0.067, p = 2.4×10^-7^).

Colocalization with cardiac expression QTLs showed uniformly weak evidence (all PP.H4<0.2), indicating these proteins act as circulating factors rather than through local cardiac gene regulation. We prioritized ECM1, CTSS, LRRC37A2, FKBPL, WNT9A, RNF5, DPY30, and APOH for external validation based on colocalization strength, effect magnitudes, and mechanistic plausibility.

### Pleiotropy testing and external validation establish CTSS and ECM1 as robust causal protein targets

To assess robustness against horizontal pleiotropy, we applied MR-PRESSO to all significant protein-phenotype pairs for the eight prioritized proteins **(Table S24)**. All associations remained valid after pleiotropy testing. CTSS and ECM1 showed global heterogeneity (RSS=135.96 and 130.77, both p = 2.0×10^-4^) with identified outliers; after outlier removal, both causal estimates remained genome-wide significant (CTSS: β = 2.78, p = 6.4×10^-6^; ECM1: β = -1.96, p = 1.2×10^-3^) with no distortion (both distortion p>0.62). FKBPL, RNF5, WNT9A, LRRC37A2, and APOH showed no evidence of pleiotropy (all global p > 0.09), with consistent effect estimates across all phenotypes.

We validated these findings by replicating the Mendelian randomization analysis in an independent Icelandic proteomics dataset (deCODE genetics, n = 35,559) **(Table S25-26)**. FKBPL, WNT9A, and RNF5 did not have available data, and LRRC37A2 lacked genome-wide significant cis-pQTLs in Iceland, consistent with prior findings that this protein cannot be robustly instrumented in deCODE^48^. ECM1 and CTSS showed robust cross-population replication, with consistent effect directions. ECM1 replicated its effect on T1 99th percentile (p = 6.2×10^-8^, 51 instruments), while CTSS demonstrated the strongest validation, replicating effects on both T1 99th percentile (p = 1.2×10^-15^, 20 instruments) and T1 standard deviation (p = 5.7×10^-12^, 20 instruments). Notably, CTSS additionally showed significant associations across VAE latent dimensions 4, 6, 8, and 9 in the Icelandic data, suggesting that cathepsin S influences both the distribution and spatial organization of myocardial fibrosis across genetically distinct European populations.

## Discussion

This study augments the phenotypic characterization of myocardial fibrosis by applying convolutional variational autoencoders (VAE) and automated distributional analysis to native T1 maps in more than 40,000 individuals **(Figure 2)**. Through genome-wide and proteome-wide association studies coupled with Mendelian randomization, we identify heritable pathways and causal circulating proteins influencing myocardial fibrosis, revealing potential therapeutic targets.

The hazard ratios observed (0.40–0.67 for protective and 1.46–2.32 for risk features) demonstrate that both spatial and distributional T1 phenotypes provide clinically meaningful risk stratification. These effect sizes modestly exceed those from prior UK Biobank analyses (HR 1.24–1.40)^49,50^. The VAE model’s improved mortality discrimination (C-index from 0.547 to 0.614; +12% relative gain) demonstrates that unsupervised learning can identify prognostic spatial patterns without requiring labeled training data or pre-specified regions of interest. This improvement is consistent with reported improvements in autoencoder-based survival models across imaging modalities (C-index gains 0.03–0.12) and with meta-analyses of machine-learning approaches for post-infarction prognosis (12–15% over clinical baselines)^51–53^. The absolute C-index of 0.614 reflects the relatively healthy UK Biobank cohort (1.8% 10-year mortality), emphasizing that spatial fibrosis features retain prognostic power even in low-risk populations.

We further establish the heritability of T1 percentiles, previously uncharacterized in cardiac imaging. Central percentiles showed heritability comparable to or exceeding mean T1 (75th h² = 0.173; 50th h² = 0.163; 25th h² = 0.146 vs. mean h² = 0.161), and were most strongly associated with mortality (75th percentile HR = 1.60, p = 3.8×10^-4^) and ischemic heart disease (25th percentile Δrank = 0.075, p = 1.0×10^-6^). These metrics therefore provide high-power genetic instruments and may capture fibrosis processes in specific myocardial segments that mean-based measures obscure. Reduced heritability at distributional tails (h² = 0.042–0.078) likely reflects greater environmental influence, or greater noise due to a higher percentage of outliers. Although mean septal T1 is the current clinical standard, incorporating central percentile-based metrics into routine reporting could improve risk stratification and yield refined phenotypes for multi-omic discovery.

Our proteomic analyses suggested a biological distinction between processes underlying T1 distributional and VAE-derived spatial features. T1 distributional metrics primarily captured systemic metabolic perturbations, as demonstrated by their strong associations with leptin (LEP) and fatty-acid-binding protein 4 (FABP4). Leptin promotes cardiomyocyte hypertrophy and fibroblast activation via oxidative and TGF-β–dependent signaling, while FABP4 alters lipid handling and fibroblast differentiation, both driving interstitial collagen accumulation^54–56^. Additionally, the enrichment of vitamin K–dependent coagulation pathways and associations with tissue plasminogen activator (PLAT) further suggest that hemostatic balance influences myocardial remodeling through fibrinolytic regulation of matrix turnover^57,58^. Together, these findings support established mechanisms of cardiac fibrosis by demonstrating that T1 percentile metrics sensitively reflect the diffuse interstitial remodeling driven by chronic inflammation and altered metabolism.

In contrast, VAE latent dimensions encode the spatial organization of extracellular-matrix (ECM) components that accumulate with age, inflammation, and mechanical stress. Structural ECM genes, COL4A1, COL6A3, and HSPG2, highlight the role of basement-membrane integrity and collagen network organization in determining tissue architecture^59–61^. Perturbations in these components are linked to matrix disarray, impaired cellular adhesion, and increased myocardial stiffness, indicating that VAE-derived spatial features may serve as a sensitive marker of the altered tissue architecture that results from chronic myocardial injury^62–64^.

Genome-wide association analyses identified both established and novel loci associated with T1 distribution metrics, converging on oxidative-stress pathways. The strongest signals localized to SOD2 and GSS, encoding sequential enzymes in mitochondrial antioxidant defense. This genetic convergence provides compelling evidence that redox imbalance is a fundamental determinant of myocardial tissue composition. Mechanistically, glutathione depletion or GSS dysfunction activates TGF-β/Smad3 signaling and matrix metalloproteinases, promoting ventricular dilation and fibrogenesis, while augmentation of glutathione reverses fibroblast activation. The association of common genetic variants in this pathway with T1 suggests that modest lifelong oxidative stress cumulatively drives myocardial fibrosis. Additional loci in calcium handling (CAMK2D, CALU) and cytoskeletal regulation (ARHGAP27, C2ORF88) support roles for excitation–contraction coupling and structural integrity in fibrosis. The absence of genome-wide significant associations for most latent dimensions supports the interpretation that spatial patterns predominantly reflect acquired rather than inherited pathology.

Mendelian randomization integrating imaging and proteomic data identified convergent causal networks across T1 distributional and spatial traits. CTSS and ECM1 emerged among the leading causal proteins with strong colocalization (PP.H4 > 0.88) and external validation. CTSS, a lysosomal cysteine protease, promotes collagen deposition and ventricular stiffening^65^; its inhibition reduces fibrosis in preclinical models, and selective small-molecule inhibitors already exist for vascular and pulmonary fibrosis^66^. ECM1 shows protective effects in murine models, consistent with its structural role in ECM stabilization, with its deficiency leading to disordered collagen architecture that can be reversed by ECM1 restoration^67^. FKBPL emerged as a significantly associated protein with both T1 distributional metrics and VAE latent dimensions. FKBPL peptides suppress fibroblast activation and collagen deposition in 3D models and modulate angiogenesis through HSP90 complexes, highlighting it as a tractable therapeutic candidate^68,69^. Additional causal associations included APOH, which may confer vascular protection by modulating immune-coagulative crosstalk, and LRRC37A2, a previously uncharacterized leucine-rich repeat gene linked to ventricular wall morphology that may mediate cardiomyocyte–ECM alignment^70,71^.

Our cross-sectional design limits assessment of fibrosis progression or therapeutic response, and the UK Biobank’s relatively healthy demographic constrains generalizability to advanced heart failure or younger populations. Important technical considerations include that while T1 is sensitive, it has limited specificity for fibrosis and can be elevated by inflammation, edema, fat, or iron deposition. Future work should validate whether AI-derived phenotypes predict therapeutic response to antifibrotic agents and whether the spatial patterns evolve predictably during disease progression. Replication in independent imaging cohorts and mechanistic validation of causal proteins, particularly LRRC37A2 and APOH, will further substantiate these findings. For proteins with existing experimental evidence (CTSS, ECM1, FKBPL), the human genetic support provided here strengthens their prioritization for translational development.

In summary, automated phenotyping via variational autoencoders and distributional analysis transforms routine cardiac imaging into high-dimensional datasets for multi-omic integration. By extracting spatial and distributional features at population scale without manual annotation, we elucidate the molecular and spatial architecture of myocardial fibrosis, refine population risk prediction, and identify actionable therapeutic targets grounded in human biology.

## Methods

### Data Acquisition and Preprocessing from the UK Biobank

This study used data from the UK Biobank (UKBB), a prospective cohort of 502,629 individuals from the United Kingdom enrolled between 2006 and 2010 with extensive phenotyping, imaging, and multiple genomic data. Initially, approximately 9.2 million individuals aged 40–69 years living in England, Scotland, and Wales were invited to participate in the study, with a 5.4% participation rate^19^.

Cardiac MRI was performed on participants in the imaging sub-study using a standardized protocol. All images were acquired on a clinical wide-bore 1.5 Tesla scanner (MAGNETOM Aera, Syngo Platform VD13A, Siemens Healthcare). Native T1 mapping was performed within a single breath hold using the cardiac-gated Shortened Modified Look-Locker Inversion recovery (ShMOLLI) technique.

In addition to T1 mapping, the UK Biobank provided comprehensive cardiac MRI structured measurements from several sequences, including CINE three-, two-, and four-chamber views, and left ventricular outflow tracts. Metabolomic data, clinical outcomes, medication history, and disease diagnoses through ICD-10 codes were also available for these participants. As of September 2024, 50,239 individuals had both cardiac MRI measurements and ShMOLLI T1-mapping images available for analysis, with 35,160 of these participants also having matched genetic data suitable for genomic analyses. The mean age of these study participants was 63.97 ± 7.65 years, and 51.18% were males.

This research was conducted under the UK Biobank’s comprehensive ethics framework (project ID 22282).

### U-Net-based Segmentation of the Myocardium

To efficiently analyze cardiac MRI data from the UK Biobank, an automated approach for myocardial segmentation was first developed. This initial step was essential as the images were unlabeled, yet required precise identification of myocardial regions before any analysis of fibrosis characteristics could proceed. The 50,239 short-axis cardiac shMOLLI T1-mapping images available at the start of the study were acquired from the UK Biobank. In these shMOLLI images, pixel-wise T1-relaxation times have been shown to correlate closely with diffuse myocardial fibrosis; in this context, longer relaxation times are typically associated with greater collagen deposition and extracellular-matrix expansion and previous studies have demonstrated that short-axis T1 mapping provides a robust representation of whole-heart fibrosis burden^13,72,73^.

To enable automated analysis of these clinically relevant measurements, segmentation masks were manually traced for the full myocardium (n=728) and then were split into training (70%), testing (20%), and validation (10%) sets.

A standardized preprocessing pipeline ensured consistent analysis across all subjects. Pixel intensities were normalized using min-max scaling to the standard [0, 255] range and converted to 8-bit unsigned integers. All images were resized to 256 by 256 using constant-mode interpolation, ensuring consistent input dimensions for downstream analyses while maintaining the integrity of the intensity values.

A U-Net architecture was trained for automated myocardial segmentation, consisting of a contracting path with four downsampling blocks (16 to 256 filters) and a symmetric expansive path with skip connections^74^ **(Supplementary Figure 1A)**. Each block used 3×3 convolutions with batch normalization and dropout (0.1) to prevent overfitting. The model was compiled using the Adam optimizer with default learning rate parameters and binary cross-entropy as the loss function, with accuracy as the evaluation metric. Training was performed for up to 100 epochs with a batch size of 16, implementing several callbacks to optimize the training process: early stopping with a patience of 10 epochs to prevent overfitting, learning rate reduction (by a factor of 0.1) after 5 epochs without improvement (with a minimum learning rate of 0.00001), and model checkpointing to save only the best-performing model based on validation performance.

The model’s predictions provided probability values (0-1) for each pixel being part of the myocardium. After examining the distribution of raw probability values, a steep dropoff was observed at 0.9, suggesting a natural separation between myocardial and non-myocardial tissue. Thus, a threshold of 0.9 was applied to create a binary mask, then OpenCV’s contour detection was used to isolate and retain only the largest connected region, effectively removing noise and ensuring a clean myocardial segmentation of the left ventricle. The binary mask was multiplied by the original T1-mapping image to generate an image consisting of only T1 values in the myocardium.

Quality control was performed using OpenCV contour tracking to confirm whether each image contained two concentric ring-shaped contours representing the endocardial and epicardial walls of the left ventricle in a short-axis view^75,76^. Images meeting these criteria were classified as good-quality myocardial ROI images. From the original 50,239 segmentations, 42,667 images (84.9%) passed the quality control step.

From the quality-controlled myocardial regions, scalar T1 mapping metrics were extracted for each subject, including mean T1, T1 standard deviation, and T1 values at seven percentile cutoffs (1st, 5th, 25th, 50th, 75th, 95th, and 99th percentiles). To remove the confounding effects of demographic factors, all extracted T1 metrics were residualized for age and sex using linear regression prior to downstream analyses.

### Advanced Spatial Modeling of the Myocardium by Variational Autoencoders

While the segmentation approach provided robust myocardial boundaries, it did not capture the rich spatial heterogeneity of fibrosis patterns within these boundaries. To address this limitation, the second approach implemented a Convolutional Variational Autoencoder (CVAE), which can learn compact representations of the complex spatial patterns within the myocardium without requiring explicit feature engineering **(Supplemental Figure 1B)**.

For each cardiac MRI image, the quality-controlled myocardial segmentation masks from the U-Net were used to isolate the region of interest. This was accomplished by multiplying the original T1-mapping image with its corresponding binary segmentation mask, creating myocardium-only representations. All 42,667 images that passed the quality control step were used for the CVAE analysis and split into training (70%), testing (20%), and validation (10%) sets, ensuring that the model learned from reliable myocardial patterns without contamination from failed segmentations.

The architecture of the CVAE model consists of a convolutional encoder and decoder network connected by a latent space representation^77^. Mathematically, we obtain:

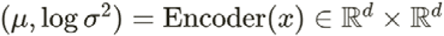

where μ and logσ^2^ represent the mean and log-variance of the latent variable distribution, and *d* is the dimension of the latent space.

We sample from this distribution using the reparameterization trick:

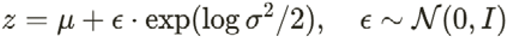

Then, the decoder reconstructs the input image:

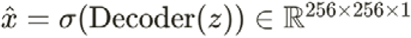

where σ is the sigmoid activation function that maps outputs to the range [0,1].

For the encoder network, a series of convolutional layers was used with increasing filter sizes (32, 64, 128, 256) and a stride of 2 for downsampling, followed by a dense layer that outputs the parameters of the latent distribution. For the decoder, a dense layer was followed by reshaping and a series of transposed convolutional layers with decreasing filter sizes (256, 128, 64, 1) and a stride of 2 for upsampling.

The loss function combines reconstruction loss with the Kullback-Leibler divergence:

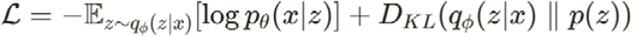

where the first term represents the negative log-likelihood of the data given the latent variable (reconstruction loss), and the second term is the KL divergence between the approximate posterior and the prior distribution. During training, this loss was optimized using the Adam optimizer with an exponential learning rate schedule starting at 10^-3^ and decaying by a factor of 0.9 every 10,000 steps.

This approach was evaluated on the task of myocardium reconstruction and representation learning, assessing the model’s ability to capture meaningful features in the latent space while accurately reconstructing the input images. This was done by calculating mean squared error (MSE), peak signal-to-noise ratio (PSNR), and the structural similarity index metric (SSIM).

Having established quantitative performance metrics for reconstruction quality, the next challenge was to interpret the learned representations in the 16-dimensional VAE model. A gradient-based attention mapping technique inspired by Grad-CAM (Gradient-weighted Class Activation Mapping) was implemented (Supplemental Figure 1B) to visualize which regions of the input images most strongly influence each individual latent dimension. For a given input image and latent dimension, this method first processes the image through the encoder network while tracking the feature maps of the final convolutional layer. Then the gradients of the specific latent dimension were computed with respect to these feature maps, capturing how changes in the activations affect the latent representation.

From these gradients, the relative importance weights for each channel were calculated in the feature maps by applying global average pooling^38^. These weights represent how strongly each channel contributes to the specific latent variable. The attention map was generated by a weighted combination of the feature maps using these importance weights, followed by applying a ReLU activation to highlight only positive contributions. The resulting attention map was then resized to match the dimensions of the original input image. This technique provides a spatial map highlighting regions in the input image that most strongly activate a particular latent dimension. By examining these maps for different latent dimensions, how the VAE decomposes and organizes different aspects of myocardial structure and tissue characteristics can be visualized.

### Prognostic and Clinical Associations of Surrogate Myocardial Fibrosis Metrics

Having developed computational methods to quantify myocardial characteristics through both scalar metrics and latent representations, the next aim was to establish their clinical relevance through a systematic analysis of their associations with patient outcomes and cardiovascular conditions **(Figure 2)**.

To assess the prognostic significance of T1 mapping metrics and VAE latent dimensions, comprehensive survival analysis was conducted. Kaplan-Meier curves were constructed to visualize the relationship between these cardiac MRI-derived metrics and all-cause mortality. The time-to-event variable was calculated as the difference between the date of cardiac MRI acquisition and the date of death or last follow-up, with patients still under observation censored at the study end date (October 18, 2022).

For each myocardial fibrosis metric (both T1 metrics and VAE latent dimensions), multiple stratification approaches were tested to identify optimal risk thresholds: median split, 25th percentile, 75th percentile, 2.5th percentile, and 97.5th percentile cutoffs. For each stratification, the study population was divided into “high” and “low” groups, and log-rank tests were performed to determine the statistical significance of survival differences between groups. Cox proportional hazards models were employed to quantify hazard ratios (HR) with 95% confidence intervals, and model discrimination was assessed using Harrell’s concordance index (C-index). As all T1 mapping metrics and VAE latent dimensions were pre-adjusted for age and sex, Cox models were fit without additional covariate adjustment.

To evaluate whether VAE-derived latent dimensions provided prognostic information beyond conventional mean T1 values, nested Cox model comparisons were performed. A baseline model containing only mean T1 was compared against a full model containing mean T1 plus all VAE latent dimensions. Model comparison was performed using likelihood ratio tests, with the test statistic calculated as 2×(log-likelihood_full - log-likelihood_baseline) and evaluated against a chi-square distribution. Model fit was additionally assessed using Akaike Information Criterion (AIC)^78^. Individual nested model comparisons were also conducted for each VAE latent dimension separately to identify which specific dimensions contributed independent prognostic information beyond mean T1 alone.

To examine whether the derived myocardial fibrosis metrics were associated with common cardiovascular comorbidities, delta rank analysis was performed. This approach evaluated the differences in metric values between individuals with and without specific conditions, focusing on several key cardiovascular and metabolic comorbidities including hypertrophic cardiomyopathy (HCM), cardiac amyloidosis, Type 1 diabetes, Type 2 diabetes, myocardial infarction, chronic kidney disease, and hypertension, as these represent common conditions with established relationships to myocardial fibrosis.

Disease phenotypes were identified using ICD-10 codes from the UK Biobank hospital episode statistics and primary care data.Specifically, the following diagnoses were extracted using their respective ICD-10 codes: type 1 diabetes (E10), type 2 diabetes (E11), myocardial infarction (I21), hypertrophic cardiomyopathy (I42.1, I42.2), dilated cardiomyopathy (I42.0), valvular disease (I05-I08, I34-I39), amyloidosis (E85), sarcoidosis (D86), chronic kidney disease (N18), hypertension (I10), and ischemic heart disease (I20-I25). Non-ischemic heart disease was defined as the presence of structural heart disease (I42-I43, I50-I51) in the absence of ischemic heart disease codes. A “Normal” reference group was defined as individuals without any ICD 10 codes. Patients were classified as having a specific disease if their medical records contained the corresponding ICD-10 code at any point in their clinical history.

For each fibrosis metric and disease pair, the median value of the metric was calculated in patients with the disease and compared it to the median value in patients without the disease. For example, when analyzing hypertension, the median T1 mapping values between the hypertension group (n=6,304) and the non-hypertension group (n=33,198) were compared. Standardized effect sizes (Cohen’s d) were calculated to enable comparison of associations across different metrics and diseases. This approach allowed quantification of the direction and magnitude of differences in fibrosis metrics associated with specific cardiovascular comorbidities.

To further characterize the relationship between the fibrosis metrics and cardiovascular comorbidities, disease prevalence across quartiles of each metric was examined. For this analysis, the study population was divided into four equal quartiles based on each fibrosis metric value: Q1 (lowest values, 0-25th percentile), Q2 (25th-50th percentile), Q3 (50th-75th percentile), and Q4 (highest values, 75th-100th percentile).

For each comorbidity of interest, the prevalence within each quartile was calculated as the percentage of individuals within that quartile who had the disease. This approach allowed for the assessment of differences in disease prevalence across the spectrum of fibrosis metric values.

To determine the statistical significance of prevalence trends across quartiles, the chi squared test was performed for each disease-metric pair. This test was chosen because it specifically assesses whether there is a linear trend in proportions across ordered categories (quartiles), making it more powerful than omnibus tests for detecting monotonic dose-response relationships. The test evaluates whether disease prevalence systematically increases or decreases across ascending quartiles of each metric. The direction of any associations was determined by comparing prevalence between the lowest (Q1) and highest (Q4) quartiles, as the Cochran-Armitage test indicates the presence but not the direction of a trend.

All statistical analyses were performed using Python 3.9.7 with NumPy (version 1.22.4), pandas (version 2.3.2), SciPy (version 1.7.1), and lifelines (version 0.30.0) packages. For each analysis method (survival analysis, delta rank analysis, and disease prevalence analysis), Bonferroni correction was applied separately to control for multiple comparisons. P-values less than 0.05 after Bonferroni correction were considered statistically significant.

### Correlation Analysis with Cardiac Structure, Metabolic Markers, and Proteomics

To investigate the relationships between T1 relaxation time metrics and metabolomic and cardiac phenotype measurements, correlation analysis was performed using the Pearson method to capture linear relationships **(Figure 2)**. The Bonferroni correction was applied to account for multiple hypothesis testing by adjusting the significance threshold to α = 0.05/n, where n represents the number of statistical tests performed. Correlations with adjusted p-values below this threshold were considered statistically significant.

The correlations between surrogate fibrosis markers (T1 scalar metrics and VAE latent dimensions) and cardiac phenotypes were evaluated including: average heart rate, ascending and descending aorta maximum and minimum areas, LV circumferential strain across all 16 American Heart Association (AHA) segments, LV longitudinal strain across six segments, LV mean myocardial wall thickness for all 16 AHA segments and global measurement, LV radial strain across all 16 AHA segments, left and right ventricular volumes (end-diastolic and end-systolic), stroke volumes, and ejection fractions, left and right atrial maximum and minimum volumes, cardiac output, cardiac index, aortic flow measurements with background correction, end-systolic circumferential strain and torsion, mean area measurements of the ascending and descending aorta during systole and diastole, pericardial fat measurements, main pulmonary artery metrics, and various standard deviation measurements of aortic areas. These cardiac phenotypes were provided directly as structural variables by the UK Biobank.

The analysis also examined relationships of surrogate fibrosis markers with metabolomic data, including apolipoprotein levels, lipid profiles (HDL, LDL, Lipoprotein A), inflammatory markers (C-reactive protein), and glycemic indices (HbA1c, glucose).

A proteome-wide association study (PWAS) was conducted utilizing proteomic profiles from the OLINK platform covering approximately 3,000 proteins as potential biomarkers related to cardiac phenotypes^79^. Ordinary least squares (OLS) regression models were implemented using the statsmodels Python package (version 0.12.2) for each combination of protein and surrogate fibrosis marker. Each protein was treated as an independent variable, with each cardiac phenotype serving as the dependent variable. The analysis focused on extracting two key parameters: beta coefficients (representing the strength and direction of association) and p-values (indicating statistical significance). To ensure data quality, regression was only performed on pairs with sufficient non-missing data points. Additionally, protein-protein interaction network analysis was conducted using STRING-db to identify functional relationships between significantly associated proteins^80^.

### Genome-wide Association Study (GWAS) of Surrogate Fibrosis Metrics

After characterizing the clinical and biochemical correlates of these fibrosis metrics, the next goal was to better understand the heritable components of myocardial tissue characteristics and potentially identify novel genetic underpinnings. To identify specific genetic loci associated with imaging-derived fibrosis phenotypes, genome-wide association studies (GWAS) were conducted to serve as the foundation for downstream genetic analyses (Figure 2).

Participants in the UK Biobank were genotyped using custom arrays, either the Affymetrix UK BiLEVE Axiom array or the Affymetrix UK Biobank Axiom array, which share 95% of marker content. The genetic data were centrally imputed into the Haplotype Reference Consortium panel and the UK10K + 1000 Genomes panel. Variant positions were identified using the GRCh37 human genome reference. Only autosomes were included in the analysis.

Only participants with both ShMOLLI T1-mapping MRI sequences and genotyping data were considered for analysis (n = 35,160). Standard quality control procedures were applied to the phenotypic data, with outlier values more than three interquartile ranges above the first quartile or below the third quartile being winsorized to minimize noise. Residuals from the phenotype variables were computed after fitting linear regression models with covariates including age, sex, BMI, and the first five principal components of population structure.

The following genetic quality control criteria were applied: filtering variants with minor allele frequency less than 1%, filtering variants with minimal allele count less than 20, excluding variants with missing call rates exceeding 1%, and excluding variants failing a Hardy-Weinberg equilibrium exact test at 1 × 10^-6^. A genome-wide association analysis of the T1 relaxation time metrics was performed using PLINK version 2.0, a general linear model (GLM)-based software package designed for whole-genome association analysis. Covariates included demographics (age, sex, BMI) and population structure (the first five principal components)^43^. PLINK was specifically selected for its computational efficiency in handling large genomic datasets and its established reliability in GWAS applications.

To adjust for multiple testing, the genome-wide significance threshold of p < 5 × 10^-8^ was applied to select genetic variants that were significantly associated with the phenotypes.

To understand the genetic architecture underlying the imaging phenotypes and quantify their genetic basis, linkage disequilibrium score regression (LDSC) analysis was performed to estimate the SNP-based heritability (h²) of the T1 mapping metrics and to quantify genetic correlations between different phenotype groups. For the LDSC analysis, the variation in each T1 mapping metric attributed to genetic factors (SNP-based heritability) was estimated by analyzing patterns in the GWAS association statistics. European ancestry reference data from the 1000 Genomes project were used for these calculations, consistent with the predominantly European ancestry of the study population.

### Causal Inference of Surrogate Fibrosis Metrics

To move beyond purely associative relationships and investigate potential causal mechanisms underlying myocardial fibrosis, Mendelian Randomization (MR) was conducted to assess potential causal relationships between protein expression levels and the imaging-derived phenotypes. Mendelian Randomization is a statistical approach that uses genetic variants as “natural experiments” to determine if an exposure (in this case, protein levels) truly causes an outcome (fibrosis metrics)^32^. Since genetic variants are inherited randomly and fixed at birth, they are not affected by the disease process or confounding factors, allowing us to establish the direction of causality. The MR workflow was implemented using the TwoSampleMR (version 0.6.14) and MendelianRandomization (version 0.10.0) R packages, and protein quantitative trait loci (pQTLs) as instrumental variables for protein expression levels^81^.

Genetic variants that influence protein levels (known as cis-protein quantitative trait loci or cis-pQTLs) were used as the exposure data. These genetic variants were located within 100 kilobases of the protein-coding gene, a region defined as “cis” in this context. To ensure the genetic instruments were independent from each other, linkage disequilibrium (LD) clumping was performed to retain only variants with minimal correlation (r² < 0.01). LD clumping identified index variants with p < 5×10^-5^ as independent lead signals, and grouped together with each index variant were any correlated variants (in LD) that had p < 1×10^-3^, forming distinct clumps of associated variants. GWAS summary statistics from latent space dimensions derived from T1 mapping and the scalar T1 mapping metrics (mean T1, T1 standard deviation, and seven additional percentile cutoffs) was used as the outcome data. As previously mentioned, data for GWAS analysis was quality controlled and regressed to age, sex, and five principal components, to meet assumptions necessary for MR. All phenotype data was residualized and quantile-normalized prior to analysis. The inverse-variance weighted (IVW) method with fixed effects was employed as the primary MR approach to estimate causal effects. The number of SNPs used as instruments was recorded for each protein-outcome pair. Results were compiled into a comprehensive table including exposure and outcome identifiers, IVW point estimates, standard errors, p-values, and the number of instrumental variables used.

Results were considered statistically significant if they achieved a p-value below the Bonferroni-corrected threshold (0.05 divided by the number of tests performed within each analysis type).

### Colocalization Analysis of Surrogate Fibrosis Metrics

To distinguish whether Mendelian randomization (MR) associations reflected true causal relationships or confounding by linkage disequilibrium, Bayesian colocalization analysis was performed using the coloc R package (version 5.2.3). This analysis tested whether protein quantitative trait loci (pQTL) signals and myocardial fibrosis GWAS signals shared the same causal variant at a given genomic locus, as opposed to being driven by two distinct but correlated variants in LD.

Colocalization analysis was restricted to protein-trait pairs that demonstrated statistically significant causal associations in the MR analysis (inverse variance-weighted p-value < 5×10^-5^). For each significant MR association, summary statistics from the corresponding cis-pQTL region and the myocardial fibrosis GWAS were extracted and harmonized.

The Bayesian colocalization method (coloc.abf) evaluates five mutually exclusive hypotheses: H0 (no association in either dataset), H1 (association only in dataset 1), H2 (association only in dataset 2), H3 (association in both datasets but with distinct causal variants), and H4 (association in both datasets driven by a shared causal variant). Only loci with at least 20 shared SNPs between the pQTL and GWAS datasets were included in the colocalization analysis to ensure sufficient statistical power.

The primary output metric was the posterior probability of hypothesis 4 (PP.H4), which represents the probability that the pQTL and GWAS signals share the same causal variant. PP.H4 > 0.8 was used as evidence for colocalization, indicating strong support for a shared causal variant. The posterior probability of hypothesis 3 (PP.H3) was also reported, representing the probability of two independent causal variants in LD. The analysis was parallelized across MR-significant protein-trait pairs using SLURM array jobs to enable efficient computation.

### Pleiotropy Analysis of Surrogate Fibrosis Metrics

The exclusivity assumption in Mendelian randomization is critical, that genetic instruments influence the outcome only through the exposure and not via alternative biological pathways. To assess potential violations, horizontal pleiotropy was evaluated using the MR-PRESSO (Mendelian Randomization Pleiotropy RESidual Sum and Outlier) framework implemented in the MRPRESSO R package (version 1.0).

MR-PRESSO analysis was applied to proteins prioritized based on significant MR associations and biological relevance to myocardial fibrosis pathways. For each protein-trait pair tested in the MR analysis, exposure (pQTL) and outcome (myocardial fibrosis GWAS) summary statistics were harmonized by aligning effect alleles. SNPs were matched between datasets, and effect sizes for the outcome were flipped when necessary to ensure alignment with the exposure allele. Allele harmonization was performed by comparing effect alleles (A1) between exposure and outcome datasets, with outcome beta coefficients inverted when alleles were misaligned.

MR-PRESSO performs three complementary tests to detect and correct for horizontal pleiotropy. The global test evaluates overall horizontal pleiotropy by comparing the observed residual sum of squares (RSSobs) to an expected null distribution generated through permutation, providing a global p-value for the presence of pleiotropy across all instruments. The outlier test identifies individual SNPs that contribute disproportionately to heterogeneity, flagging variants as outliers based on a significance threshold of p < 0.05. The distortion test evaluates whether removal of identified outliers significantly alters the causal estimate, quantified by a distortion coefficient and associated p-value.

For each protein-trait pair, MR-PRESSO was run with 5,000 permutations to generate robust null distributions for hypothesis testing. The analysis produced both raw causal estimates (including all instruments) and outlier-corrected causal estimates (excluding identified pleiotropic variants). Pairs with fewer than three shared SNPs were excluded from MR-PRESSO analysis due to insufficient statistical power.

A significant global test (p < 0.05) indicates the presence of horizontal pleiotropy across the instrument set; identified outlier SNPs represent individual variants likely acting through pleiotropic pathways; and a significant distortion test (p < 0.05) suggests that pleiotropic outliers meaningfully bias the causal estimate, warranting use of the outlier-corrected estimate for inference. Associations with non-significant distortion tests were considered robust to pleiotropy, supporting the validity of the causal inference.

### External Validation in Independent Cohort

To assess the generalizability and robustness of MR findings across genetically distinct populations, external replication was performed using the deCODE genetics proteomics GWAS dataset comprising 35,559 Icelandic participants. Replication was attempted for proteins prioritized in the primary analysis that had available proteomics data in deCODE. The same MR analytical framework described above was applied to proteins with available deCODE pQTL data. For each protein with genome-wide significant cis-pQTLs in the deCODE dataset, genetic instruments were extracted and harmonized with the UK Biobank myocardial fibrosis GWAS outcomes. The inverse-variance weighted method was used to estimate causal effects, and results were considered replicated if they showed consistent effect directions and achieved nominal significance (p < 0.05). Proteins lacking genome-wide significant cis-pQTLs in the Icelandic population or without available proteomics data in deCODE were noted as unable to be instrumented for replication analysis.

## Data Availability

Access to UK Biobank data is available to approved researchers via https://www.ukbiobank.ac.uk/. Summary statistics and analysis results from this study are in the supplemental data or available upon reasonable request to the authors.

https://docs.google.com/spreadsheets/d/14UgoXGtmIcO1ipOANF4-ABZr2d9963MTRXw3Q8MQbvQ/edit?gid=1688534539#gid=1688534539

## Notes

### Competing Interest Statement

E.A. is founder of Personalis, DeepCell, Svexa; advisor to Pacific Biosciences, SequencBio, and Apple and non-executive director of AstraZeneca. J.S. is a Consultant for Google AI. V.P. is SAB for BioMarin and Lexeo Therapeutics; a Consultant for viz.ai and Nuevocor and has sponsored research from Biomarin, Inc. and Saliogen Therapeutics. M.W is a Consultant for Cytokinetics and and has sponsored research from Astra Zeneca, Bristol Myers Squib, Salubris Bio and Cytokinetics, Nikon Kohden. All other authors have no conflicts of interest to declare.

### Funding Statement

This research was supported by the AHA postdoctoral fellowship - 23POST1019783

### Author Declarations

UK Biobank received ethical approval from the National Health Service's National Research Ethics Service North West (11/NW/0382), and this research was conducted under UK Biobank application 22282.

