## Supplemental Data for "Deep learning representations and proteome-wide Mendelian randomization identify causal mediators of myocardial fibrosis"

### Supplementary Figures

**Supplemental Figure 1: Deep Learning Architecture for Myocardial Segmentation and Spatial Feature Extraction** (A) Myocardial segmentation U-Net architecture showing the contracting and expansive paths with skip connections, illustrating filter sizes at each level from input to output. (B) Variational autoencoder architecture and attention map pipeline demonstrating the encoding of T1-relaxation maps into a latent space and the gradient-based attention map visualization approach^38^.

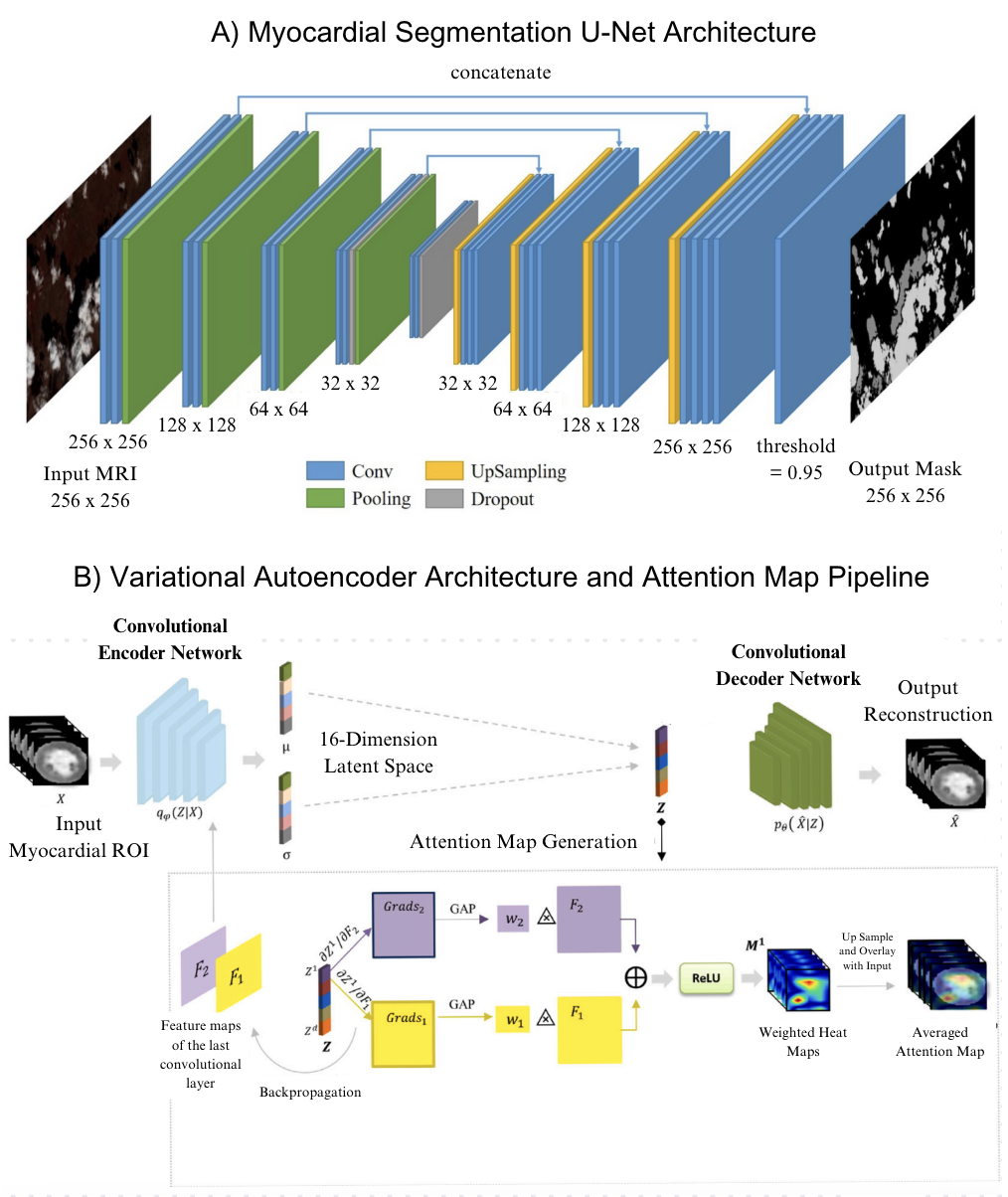

**Supplemental Figure 2: Optimal Latent Space Dimensionality for Capturing Myocardial T1 Map Features.** Performance metrics for different latent space dimensionalities show that 16 dimensions represent an optimal trade-off point. A) Mean squared error (MSE) decreases substantially until 16 dimensions, while peak signal-to-noise ratio (PSNR) reaches the quality threshold. Bottom: Metrics including SSIM, reconstruction impact, perturbation sensitivity, and KL divergence converge at 16 dimensions. B) Visualization showing how latent spaces represent spatial components of T1 relaxation maps across septum thickness and sphericity.

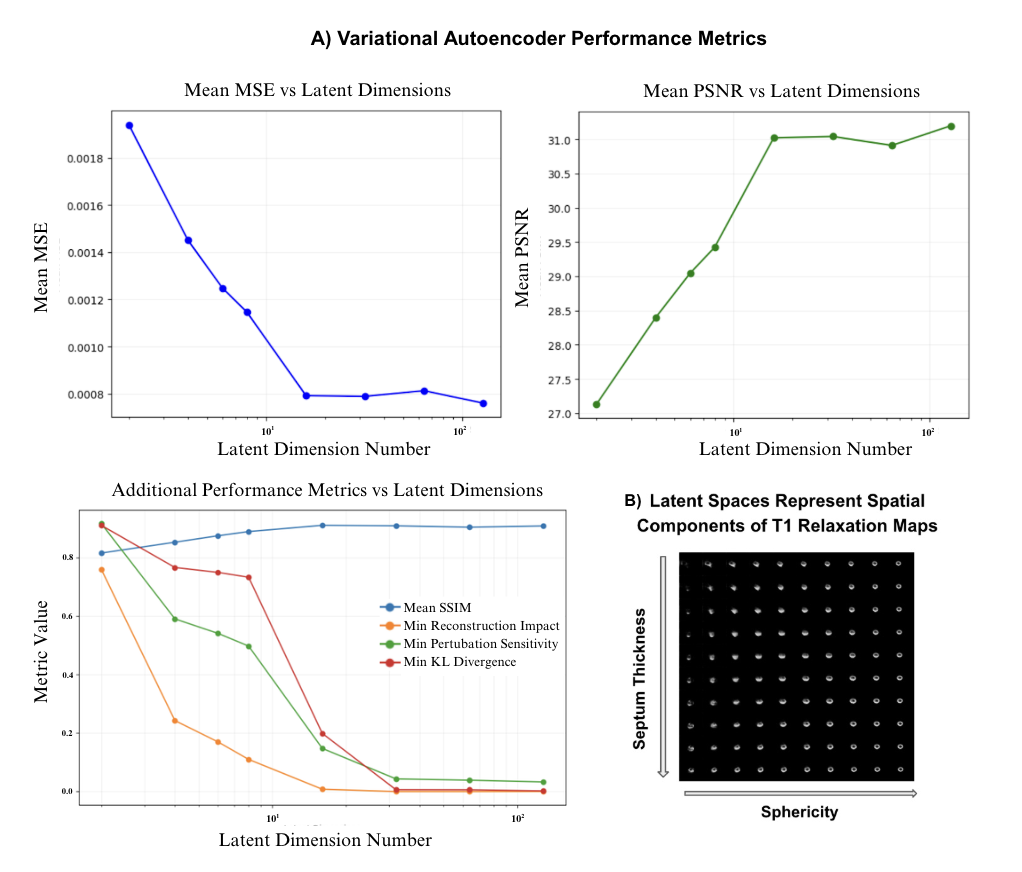

**Supplementary Figure 3: U-NET segmentation validation for automated myocardial T1 mapping** A) Automated segmentation pipeline showing input shMOLLI image, U-NET prediction, post-processing, and comparison with ground truth (Dice = 0.920). Yellow and cyan contours show predicted and ground truth myocardial masks, respectively, with extracted mean T1 = 898 ms. B) Bland-Altman analysis showing excellent agreement between predicted and ground truth T1 measurements (mean difference: -1.88 ms; 95% limits of agreement: -35.25 to 31.49 ms; n = 105). C) Strong correlation between automated and expert T1 quantification (r = 0.956, R² = 0.914, p = 1.38×10⁻⁵⁶, n = 105), validating the accuracy of automated T1 distribution analysis.

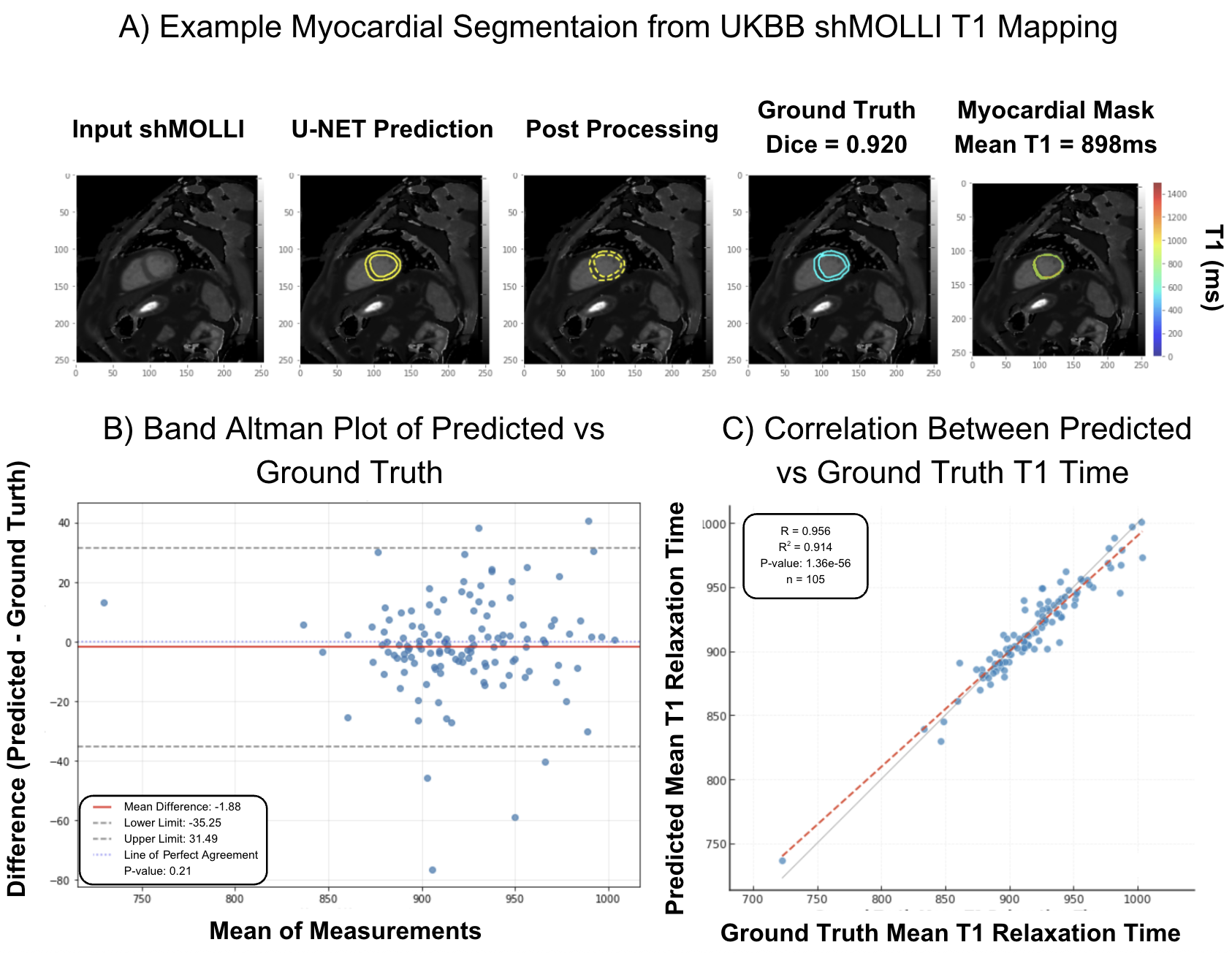

**Supplemental Figure 4: Delta Rank Analysis of T1 Mapping Metrics and VAE Latent Dimensions Across Cardiovascular Conditions.** The standardized effect sizes of associations between various cardiovascular conditions and both A) conventional T1 mapping metrics (left panels) and B) VAE latent dimensions (right panels). Higher values indicate stronger associations between the condition and the corresponding metric. Statistical significance after Bonferroni correction (p < 0.00025) is indicated with asterisks. Significant associations were observed for Hypertension, Type 2 Diabetes, and Myocardial Infarction, with distinct patterns of association with conventional metrics versus VAE latent dimensions.

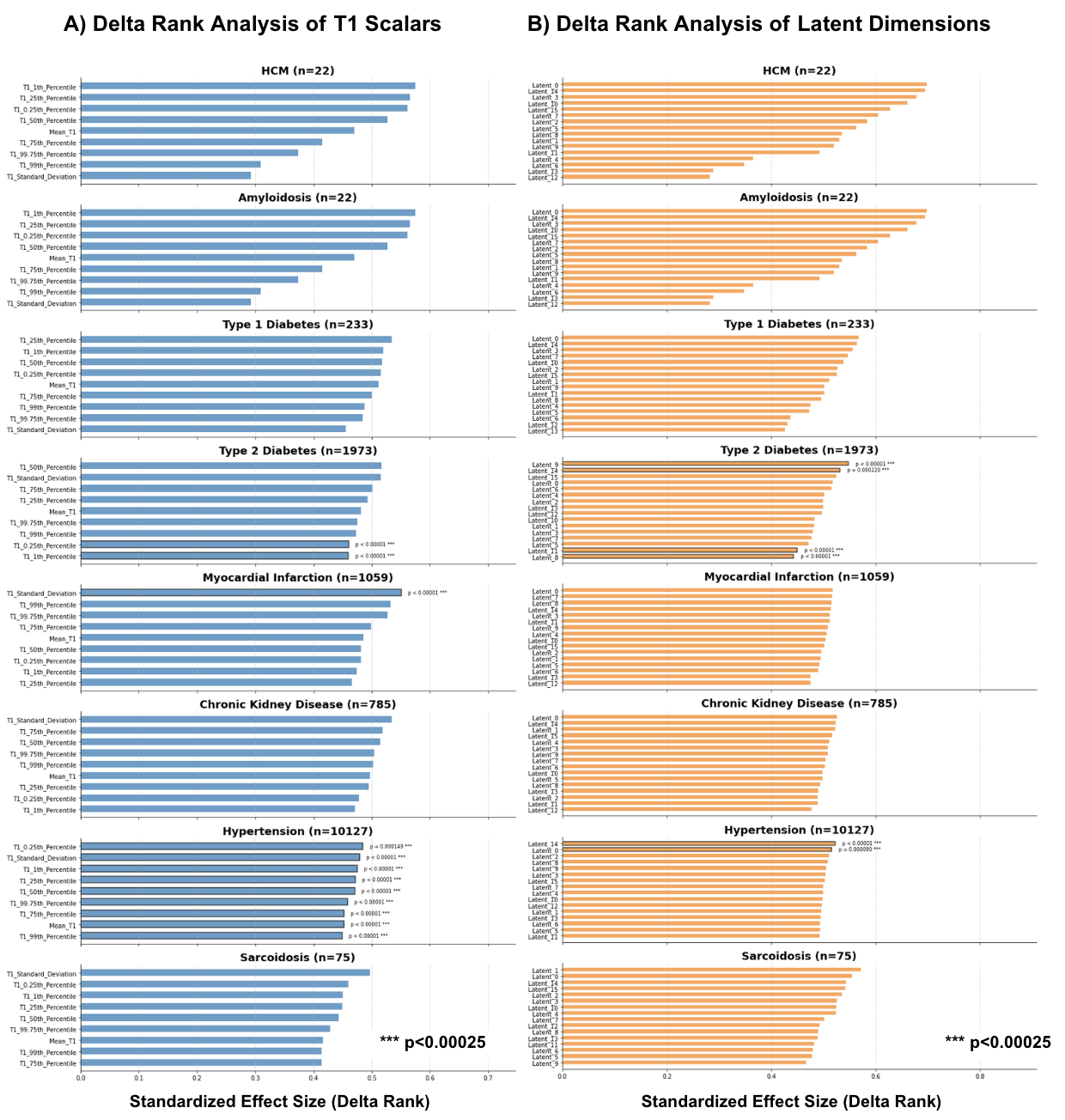

### **Supplementary Tables**

[Supplementary Tables: Deep learning representations and proteome-wide Mendelian randomization identify causal mediators of myocardial fibrosis](https://docs.google.com/spreadsheets/d/14UgoXGtmIcO1ipOANF4-ABZr2d9963MTRXw3Q8MQbvQ/edit?gid=1688534539#gid=1688534539)

##### I. Descriptive Statistics and Phenotype Characterization

**Table S1:** Summary statistics for T1 distribution metrics, stratified by sex

**Table S2:** Summary statistics for VAE-derived latent dimensions

**Table S3:** Disease prevalence stratified by cardiac imaging phenotype quartiles

##### II. Survival and Clinical Outcomes

**Table S4:** UK Biobank cohort characteristics and follow-up for survival analysis

**Table S5:** Mortality risk stratification by percentile thresholds of T1 and VAE imaging features: Kaplan-Meier and Cox regression results

**Table S6:** Nested Cox model comparison: Mean T1 vs. Mean T1 + VAE latent dimensions

**Table S7:** Incremental predictive value of individual VAE latent dimensions through nested model comparisons

##### III. Phenotypic Correlations

**Table S8:** Significant correlations between U-Net T1 mapping measures and VAE-derived cardiac phenotypes (Bonferroni-adjusted)

**Table S9:** Significant correlations between U-Net T1 mapping measures and metabolomic measures (Bonferroni-adjusted)

**Table S10:** Significant correlations between U-Net T1 mapping measures and cardiac phenotypes (Bonferroni-adjusted)

**Table S11:** Significant correlations between VAE latent dimensions and cardiac phenotypes (Bonferroni-adjusted)

**Table S12:** Significant correlations between VAE latent dimensions and metabolomic measures (Bonferroni-adjusted)

##### IV. Proteome-Wide Association Studies (PWAS)

**Table S13:** Significant plasma protein associations with T1 mapping percentiles and distribution metrics (age-, sex-, and household-adjusted)

**Table S14:** Significant plasma protein associations with VAE-derived cardiac phenotypes (age- and sex-adjusted)

**Table S15:** Enriched Reactome pathways for proteins significantly associated with myocardial T1 mapping percentiles (STRING analysis)

**Table S16:** Enriched Reactome pathways for proteins significantly associated with VAE-derived cardiac phenotypes (STRING analysis)

##### V. Genetic Architecture

**Table S17:** SNP-based heritability (h²) estimates for myocardial T1 mapping distribution metrics

**Table S18:** SNP-based heritability (h²) estimates for VAE-derived latent cardiac dimensions

**Table S19:** Genome-wide significant loci for T1 mapping phenotypes (p < 5×10⁻⁸)

##### VI. Causal Inference: Mendelian Randomization and Validation

**Table S20:** Mendelian randomization analysis of plasma proteins and T1 mapping distribution metrics

**Table S21:** Mendelian randomization analysis of plasma proteins and VAE-derived cardiac phenotypes

**Table S22:** Bayesian colocalization analysis of protein QTLs and T1 mapping GWAS signals

**Table S23:** Bayesian colocalization analysis of protein QTLs and VAE latent dimension GWAS signals

**Table S24:** MR-PRESSO pleiotropy testing for prioritized protein-phenotype associations

**Table S25:** External replication of protein-T1 distribution metric associations in deCODE Genetics cohort (n=35,559)

**Table S26:** External replication of protein-VAE associations in deCODE Genetics cohort (n=35,559)

### **Supplementary Notes**

#### Current Knowledge of Fibrosis Development Pathways and Treatments

Cardiac fibrosis represents a complex pathophysiological process with multiple overlapping signaling pathways contributing to its development and progression. At its core, fibrosis progression involves myofibroblast activation through several key pathways, including transforming growth factor-β (TGF-β), Wnt, and Notch signaling, which collectively drive epithelial/endothelial-mesenchymal transition (EMT/EndMT) and excessive extracellular matrix (ECM) deposition^6,7,8^. While TGF-β signaling remains central to fibrosis development across multiple organ systems, the redundancy created by overlapping pathways (Wnt, PI3K/AKT, JAK/STAT) presents a significant challenge for targeted therapeutic interventions^9^. This signaling complexity is further compounded by myofibroblast resistance to apoptosis, which enables these cells to persist and continue ECM production even when initial activating signals have diminished^10^.

The spatiotemporal complexity of cardiac fibrosis progression further complicates therapeutic development. Fibrosis involves dynamic cell-ECM interactions where mechanical stiffness propagates fibroblast activation independently of soluble factors, creating self-sustaining feedback loops that become increasingly treatment-resistant over time^10^. Established anti-fibrotics like nintedanib demonstrate limited efficacy against ECM stiffness, highlighting how current therapeutics merely slow rather than reverse fibrotic processes. Due to the complicated interaction of multiple signaling pathways in cardiac fibrosis development, multitarget drug regimens would likely be beneficial for effective therapy, an approach that requires better characterization of underlying molecular mechanisms.

While LGE remains the gold standard for fibrosis quantification, its need for an invasive contrast agent impedes the accumulation of large-scale image databases. Fully non-invasive MRI-based techniques enable the collection of substantially larger datasets with enhanced statistical power. These approaches can capture both the extent and spatial distribution of fibrosis and, when integrated with genomic and proteomic data, offer a promising path to identify novel therapeutic targets that address the complex, self-reinforcing nature of the fibrotic process in heart failure.

#### Cardiac Fibrosis Measurement in Imaging

Recent advancements in cMRI, particularly T1-relaxation mapping methods such as Shortened Modified Look-Locker Inversion Recovery (shMOLLI), offer a contrast-free alternative to LGE with greater sensitivity in detecting diffuse myocardial fibrosis^5^. T1 relaxation time is an MRI parameter that measures the time taken for protons to realign with the external magnetic field after being excited by a radiofrequency pulse. When fibrosis occurs, the extracellular matrix expands due to collagen deposition, altering the molecular environment and proton interactions within the tissue, which results in a longer T1 relaxation time. ShMOLLI has emerged as a robust, quantitative technique for tissue characterization that can detect early fibrotic changes often missed by conventional LGE imaging^11,12^. Validation studies comparing shMOLLI against histological samples have demonstrated strong correlations between elevated T1 times and increased extracellular volume fraction, a hallmark of fibrosis^13,14,15^.

It is important to note, however, that T1 relaxation time can be influenced by other factors beyond fibrosis, including edema, inflammation, and iron deposition, which necessitates careful interpretation of T1 mapping data^16,17,18^. Nevertheless, T1 mapping is widely adopted clinically, providing standardized quantitative values that allow for comparative assessment across different centers and patients, making it ideal for large-scale population studies.

#### Spatial Pattern Recognition in Cardiac Imaging

Recognizing that spatial context often holds the key, the integration of spatial information about fibrotic distribution is critical for uncovering genomic and molecular insights into various diseases. Different fibrotic patterns manifest in distinct ways across diseases and disease stages, providing valuable clues about underlying pathophysiological mechanisms (Figure 1). For instance, in idiopathic pulmonary fibrosis, a honeycomb pattern of fibrosis is characteristic, while in nonspecific interstitial pneumonia, a more diffuse ground-glass appearance is typical^23,24^. These spatial patterns not only aid in diagnosis but also offer insights into disease progression and potential therapeutic targets.

To capture such patterns computationally, variational autoencoders (VAEs) have emerged as powerful tools across neuroimaging, radiology, and histopathology for capturing complex spatial patterns in a low-dimensional latent space while simultaneously preserving anatomical features and extracting clinically relevant biomarkers without explicit annotation^25-28^. Unlike traditional dimensionality-reduction techniques, VAEs learn a probabilistic mapping between the input space and latent variables, ensuring that similar images cluster together in the latent space while maintaining meaningful variation.

A key advantage of VAEs for cardiac imaging analysis is their ability to integrate both tissue characterization parameters (such as T1 surrogates) and morphological features into a unified representation, potentially capturing interactions between fibrosis patterns and cardiac structure. Recent advances in attention mechanisms have further enhanced VAEs' ability to focus on regions of interest within images, making them particularly suitable for identifying spatially heterogeneous patterns such as fibrosis distribution across the myocardium^29,30^. Bonazzola et al. demonstrated that unsupervised VAEs could identify 49 loci influencing left ventricular morphology from 3D meshes, including TTN variants associated with fibrosis-prone geometries, highlighting the genetic underpinnings of structural and functional cardiac phenotypes^27^. In the same study, VAEs boosted the discovery of SMARCB1 variants influencing both fibrosis and trabeculation through shared latent pathways.

This evidence positions VAEs as critical tools for decoding the triad of spatial fibrosis patterns, proteomic signatures, and genetic architecture in population imaging.

#### Technical Validation of Myocardial Segmentation

Building on established approaches that extract mean septal T1^20^, we developed an automated pipeline to segment the entire left ventricular myocardium and quantify the full distribution of T1 values within a short-axis slice of each heart. We analyzed 50,239 UK Biobank participants (mean age 64.0±7.7 years, 48.6% male) who underwent native T1 mapping using the Shortened Modified Look-Locker Inversion recovery (ShMOLLI) sequence, a contrast-free technique that measures myocardial tissue relaxation times to quantify fibrosis^12,35^.

The approach worked well at scale. The U-Net models demonstrated robust performance **(Figure 3A)** with a Sørensen–Dice score of 0.84 (95% CI: 0.83–0.85) for myocardial segmentation on quality-controlled test images (n=105), comparable to or exceeding previous work by Nauffal *et al*. (Sørensen–Dice coefficient = 0.82 for interventricular septum segmentation, 95% CI = 0.70–0.94)^19^. Very high consistency was found between automated and manual measurements (paired t-test: t=1.745, p=0.084), with a mean difference of 1.96 ms (95% limits of agreement: −20.6 to 24.5 ms) and strong correlation (Pearson r=0.914, p < 10^-56^). After quality control, automated myocardial segmentation succeeded in 42,083 participants (83.7% overall, with 99.5% success among the subset with available genetic data).

To systematically characterize T1 variability, we extracted multiple distribution metrics from the myocardial region in each individual T1 map: extreme percentiles (0.25th, 1st, 99th, 99.75th), quartiles (25th, 50th, 75th), mean, and standard deviation **(Figure 2, Supplementary Table X)**. Mean myocardial T1 across all subjects was 918.8 ± 33.8 ms (mean ± SD), with an interquartile range of 898.07 ms to 919.06 ms. The overall distribution demonstrated negative skewness (skewness coefficient = -0.35). Consistent with established patterns, females exhibited higher native T1 values than males^36,37^. These demographic effects were consistent across all T1 distribution metrics **(Supplementary Table X)**. Therefore, all subsequent analyses were adjusted for age and sex to isolate associations independent of these normal physiological variations. While these summary statistics capture the distribution of T1 values, they do not preserve the spatial relationships and regional patterns within the myocardium, motivating our second analytical approach.

#### Technical Validation of the Variational Autoencoder Approach

To determine the optimal latent space dimensionality for the convolutional variational autoencoder (CVAE) model, multiple configurations ranging from 2 to 128 dimensions were evaluated **(Supplemental Figure 2)**. The analysis revealed that 16 dimensions represent an optimal trade-off point across all metrics. The mean squared error (MSE) decreased substantially from 2 to 16 dimensions (from 0.0019 to 0.0008) but plateaued thereafter, while peak signal-to-noise ratio (PSNR) simultaneously reached the perceptual quality threshold of 31.0 dB. At 16 dimensions, the structural similarity index measure (SSIM) approached its maximum value (~0.92) while regularization metrics (reconstruction impact, perturbation sensitivity, and KL divergence) converged toward minimal values. This empirical evidence indicated that 16 dimensions provided sufficient capacity to encode essential cardiac morphological features while maintaining computational efficiency; therefore, this number was selected for the final implementation.

#### Mechanisms and Druggability of Top Causally Linked Protein Candidates

LRRC37A2 emerged as one of the most significant colocalizing loci across spatial fibrosis traits, mapping to a region previously associated with cardiac morphology and ventricular wall structure. Although the leucine-rich repeat–containing family has been largely uncharacterized in cardiac biology, LRRC37A2’s proximity to genes involved in cytoskeletal anchoring and ciliary signaling suggests a role in maintaining cardiomyocyte-ECM alignment. Perturbations in this axis may predispose to maladaptive remodeling and collagen accumulation. The MR evidence linking LRRC37A2 expression to increased fibrosis highlights it as a potential novel structural regulator of myocardial integrity.

Cathepsin S (CTSS), a lysosomal cysteine protease, demonstrated strong positive causal effects on fibrosis phenotypes. CTSS is highly expressed in activated cardiac fibroblasts and macrophages during matrix remodeling and inflammation. Inhibition of CTSS in preclinical models reduces collagen deposition, inflammatory infiltration, and ventricular stiffening. The MR signal here suggests that CTSS activity contributes causally to myocardial fibrosis. Given the availability of selective small-molecule CTSS inhibitors already tested in vascular and pulmonary fibrosis, repurposing these agents represents a tractable therapeutic avenue for cardiac remodeling.

Extracellular Matrix Protein 1 (ECM1) showed a strong negative association with myocardial fibrosis, consistent with its established role as a structural scaffold and regulator of ECM organization. ECM1 interacts with fibulins, laminins, and proteoglycans to stabilize basement membrane architecture, and its deficiency leads to disordered collagen assembly and dermal fibrosis. The protective MR association implies that higher circulating ECM1 levels may restrain pathological matrix deposition, possibly by buffering fibroblast activation or modulating TGF-β availability. Therapeutically, strategies that enhance ECM1 expression or mimic its stabilizing interactions could offer a novel means of normalizing ECM structure in cardiac disease.

Apolipoprotein H (APOH), better known for its roles in coagulation and immune complex clearance, exhibited significant causal effects on spatial fibrosis traits. Chronic inflammatory and pro-thrombotic states accelerate cardiac fibrosis through endothelial activation and microvascular dysfunction. APOH binds phospholipid surfaces and modulates complement activation and platelet aggregation, positioning it as a regulatory hub in the vascular–fibrotic interface. The MR findings suggest that APOH exerts a protective influence, possibly by dampening immune-coagulative crosstalk that drives fibroblast recruitment. Enhancing APOH function or mimicking its anticoagulant actions may thus represent an underexplored antifibrotic strategy targeting vascular inflammation.

FKBPL emerged as one of the most significant causal proteins across both T1 and latent traits. Recent data show that FKBPL peptides reduce fibroblast activation and collagen deposition in 3D cardiac fibrosis models. FKBPL regulates angiogenesis and stress signaling via HSP90 complexes, suggesting that enhancing FKBPL activity could restore vascular–fibrotic balance. Given its extracellular accessibility and existing small peptide derivatives, FKBPL represents a tractable anti-fibrotic target.

RNF5, an E3 ubiquitin ligase involved in ER stress and proteostasis, displayed consistent MR effects across latent phenotypes. RNF5 knockout mice develop exaggerated cardiac hypertrophy and fibrosis, and pharmacologic enhancement of RNF5-mediated degradation pathways has been proposed for cystic fibrosis. These data suggest that augmenting RNF5 function could counteract maladaptive stress responses driving cardiac fibrosis.

DPY30 showed one of the strongest negative MR associations, suggesting a protective role against fibrosis. As a core component of the SET1/MLL histone methyltransferase complex, DPY30 regulates chromatin accessibility at metabolic and fibrotic gene loci. Epigenetic activators of DPY30 or stabilizers of its complex could represent an unconventional yet powerful antifibrotic strategy.

WNT9A, strongly positively associated with latent fibrosis traits, reinforces the canonical Wnt/TGF-β axis as a key effector of fibroblast activation. Several WNT9A inhibitors are in early development for renal and pulmonary fibrosis, raising the possibility of repurposing such agents for cardiac indications.
